# Cortical iron-related markers are elevated in mild Traumatic Brain Injury: An individual-level quantitative susceptibility mapping study

**DOI:** 10.1101/2024.10.29.24316391

**Authors:** Christi A. Essex, Devon K. Overson, Jenna L. Merenstein, Trong-Kha Truong, David J. Madden, Mayan J. Bedggood, Catherine Morgan, Helen Murray, Samantha J. Holdsworth, Ashley W. Stewart, Richard L. M. Faull, Patria Hume, Alice Theadom, Mangor Pedersen

## Abstract

Quantitative susceptibility mapping (QSM) has been applied to map brain iron distribution after mild traumatic brain injury (mTBI), to understand properties of neural tissue which may be related to microstructural damage. However, mTBI is a heterogeneous injury associated with microstructural brain changes, and ‘traditional’ group-wise statistical approaches may lead to a loss of clinically relevant information, as subtle individual-level changes can be obscured by averages and confounded by within-group variability. More precise and individualised approaches are needed to characterise mTBI better and elucidate potential cellular mechanisms to improve intervention and rehabilitation. To address this issue, we build individualised profiles of regional positive (iron-related) magnetic susceptibility across 34 bilateral cortical regions of interest (ROIs) following mTBI. Healthy population templates were constructed for each cortical area using standardised z-scores derived from 25 age-matched male controls, serving as a reference against which z-scores of 35 males with acute (< 14 days) sports-related mTBI (sr-mTBI) were compared. Secondary analyses sensitive to cortical depth and curvature were also generated to approximate the location of iron accumulation in the cortical laminae and the effect of gyrification. Our primary analyses indicated that approximately one-third (11/35; 31%) of mTBI participants exhibited elevated positive susceptibility indicative of abnormal iron profiles relative to the healthy control population, a finding that was mainly concentrated in ROIs within the temporal lobe. Injury severity was significantly higher (p < 0.01) for these mTBI participants than their iron-normal counterparts, suggesting a link between injury severity, symptom burden, and elevated cortical iron. Secondary analyses of cortical depth and curvature profiles revealed abnormal iron accumulation in 83% (29/35) of mTBI participants, enabling better localisation of mTBI-related changes in iron content to specific loci within each ROI and identifying effects that may be more subtle and lost in ROI-wise averaging. Our findings suggest that individualised approaches can further elucidate the clinical relevance of iron in mTBI. Differences in injury severity between iron-normal and iron-abnormal mTBI participants highlight not only why precise investigation is required to understand the link between objective changes in the brain and subjective symptomatology, but also identify iron as a candidate biomarker for tissue damage after mTBI.

## INTRODUCTION

Exposure to mild traumatic brain injury (mTBI) is a significant public and personal health concern, accounting for approximately 90% of the 50-60 million annual cases of TBI worldwide.^1^ Global financial losses related to mTBI are estimated at ~USD $400 billion per year,^1,2^ however, beyond the economic impacts mTBI can increase the risk of neurodegeneration, dementia,^3,4^ and premature death.^5^ In the short term, mTBI can result in a range of symptoms with significant inter-individual variability, including cognitive, emotional, and physiological disturbances such as sleep disruption, light sensitivity, fatigue, headaches, vertigo, vestibular problems, depression, and anxiety, which significantly impact quality of life and participation in day-to-day activities for many.^4^ In some cases, these symptoms can persist even up to three decades post-injury.^6,7^ Numerous factors contribute to differences in injury severity, symptom burden, *in-vivo* brain tissue pathology, and even autopsy findings. These include individual differences prior to injury such as genetic predispositions, age, gender, IQ, psychiatric history, prior exposure to mTBI, and substance use history, as well as differences in the mechanisms and loci of injury.^8^ In sports-related mTBI (sr-mTBI), for example, variability in the sport and even player position can affect injury severity, lead to diverse effects on brain structure and function, and divergence in symptom burden and cluster.^8^

The heterogeneity of mTBI is apparent at even the cellular level. The rapid changes in inertia (acceleration/deceleration/rotation) or exogenous skull impact associated with mTBI cause the transmission of mechanical forces to the brain, resulting in a mechanistically-specific primary insult and microstructural tissue damage.^8,9^ This initiates a variable cascade of secondary cellular processes, including disruption of the blood-brain barrier (BBB), cerebrovascular dysfunction, oxidative stress, axonal degeneration, and neuroinflammation^9,10^ which can propagate for months after the initial impact.^11^ However, the pathophysiology of mTBI remains poorly understood, and specific biomarkers indicative of mTBI remain, to date, elusive. Unlike moderate-to-severe TBI (ms-TBI), where lesions, haemorrhages, or macroscopic morphological abnormalities can be detected, routine magnetic resonance imaging (MRI) methods are often insensitive to mTBI-related neuropathology.^12,13^ This limitation necessitates the use of advanced MRI techniques not typically employed in conventional medical settings to identify the subtle changes in brain structure characteristic of this ‘mild’ injury.^14,15^ Integrating these advanced imaging modalities into routine patient care requires further validation and the establishment of clinically and individually relevant biomarkers for mTBI diagnosis and treatment.

Iron accumulation is increasingly recognised as a component of neuropathology following mTBI, contributing not only to acute-phase secondary injury and later cell death,^9^ but also cognitive dysfunction after mTBI.^16,17^ Quantitative susceptibility mapping (QSM) is an advanced MRI technique that can be used to estimate the substrates of biological tissue by leveraging the inherent magnetic properties, such as paramagnetism exhibited by iron in response to an applied magnetic field.^18–23^ Non-heme iron (particularly ferritin-bound iron), is the main source of paramagnetism on QSM^18,24–26^ and widely recognised as the form of iron most involved in secondary injury after mTBI.^9,27,28^ Iron dyshomeostasis can trigger auto-toxic circuits that drive neurodegenerative processes,^29^ including the generation of reactive oxygen species (ROS), which at high levels can lead to cytotoxic oxidative stress,^30^ lipid damage, and increased permeability of the cell membrane,^9^ as well as iron-regulated cell death (ferroptosis).^31^ As such, elevated levels of iron in cortical regions would suggest localisation of injury-related pathological processes and changes in brain structure. These changes may be related, but not limited to, mTBI-induced permeability of the BBB^32^ and neuroinflammation,^33^ both of which are known to be involved in iron accumulation.^9,30^ Iron has also been implicated in the hyperphosphorylation of tau proteins (p-tau)^28^ observed in mTBI-related tauopathies; its co-localisation with p-tau thus identifies it as a promising early indicator of neurodegeneration.^34,35^

A limited number of studies have employed QSM to investigate the role of brain iron in microstructural tissue damage following mTBI, focusing mainly on subcortical nuclei or global grey and/or white matter,^36–44^ with only a few studies including cerebral regions of interest (ROIs)^38^ or investigating the relevance of cortical morphology.^45^ However, the diversity of mTBI effects may not be discernible at the group level, which currently constitutes the standard statistical approach. Individual-level investigations of injury-specific effects may better characterise mTBI-related neuropathology, and are increasingly recognised as providing more biologically informative data than group-level studies, especially in clinical populations where targeted interventions are both useful and necessary,^46^ such as mTBI. Personalised profiles can be generated by leveraging z-scores to compare the results of individual quantitative measures to the distribution of a healthy normative population. This approach allows for a clearer understanding of where the individual falls relative to normal ranges for selected metrics.^47^ Individual analytic approaches have been successfully applied in the context of mTBI using T2 relaxometry as a marker of neuroinflammation,^48^ and diffusion-weighted imaging to investigate white matter fiber tracts.^49^ Under the TBI umbrella more broadly, individual analyses have been applied to fixel-based analysis of diffusion MRI^50^ and diffusion tensor imaging^51^ to investigate white matter integrity, as well as structural connectomics,^52^ in ms-TBI. One study has used QSM to generate individualised profiles of iron deposition in ms-TBI^47^ however, to date, dedicated personalised investigations of iron deposition at the individual level following *mild* TBI are, to the authors knowledge, lacking.

To address these research gaps, we conducted the first dedicated individual-level investigation of iron-related mTBI effects. This study aimed to: 1) generate individual profiles of cortical iron deposition following sr-mTBI, and; 2) extend these findings by deriving profiles sensitive to cortical architectonics (depth and curvature) as a supplemental, secondary approach. Our prior research suggests that elevated magnetic susceptibility should be most evident in ROIs in the temporal lobe.^45^ However, given the preliminary and exploratory nature of this study, we did not have specific *a priori* hypotheses about the direction of effects in all cortical regions.

## MATERIALS AND METHODS

Ethical approval for this research was obtained from the Health and Disabilities Ethics Committee (HDEC) (Date: 18/02/2022, Reference: 2022 EXP 11078) and institutional approval was also obtained from the Auckland University of Technology Ethics Committee (AUTEC) (Date: 18/02/2022, Reference: 22/12). In accordance with the Declaration of Helsinki, all participants provided written informed consent prior to data collection.

### Participants

35 male contact sports players (*M* = 21.60 years [range: 16-33], *SD* = 4.98) with acute sr-mTBI (sustained within 14 days of MRI scanning (*M* = 10.40 days, *SD* = 3.01)) and 25 age-matched male controls (*M* = 21.10 years [range: 16-32], *SD* = 4.35) were recruited for this observational, case-control study (see Table 1). Ages were not significantly different between groups (*t*(58) = −0.44, *p* = .66). Clinical (sr-mTBI) participants were recruited through three Axis Sports Medicine clinics (Auckland, New Zealand), via print and social media advertisements, word-of-mouth, and through community-based pathways including referrals from healthcare professionals and sports team management. Each clinical participant was required to have a confirmed sr-mTBI diagnosis by a licensed physician as a prerequisite for study inclusion, and symptom severity was assessed using the Brain Injury Screening Tool (BIST)^53^ either upon presentation to Axis clinics or electronically following recruitment. Healthy controls (HC) were recruited through print and social media advertisements, and word-of-mouth. Exclusion criteria for all participants included a history of significant medical or neurological conditions unrelated to the study’s objectives and contraindications for MRI. Additionally, controls were excluded if they had any recent history of mTBI events (< 12 months) or were living with any long-term effects of previous mTBI. All participants completed a brief demographic questionnaire and attended a 1-hour MRI scan at The Centre for Advanced MRI (CAMRI), Auckland, New Zealand. All scans were reviewed by a certified neuro radiologist and consultant for clinically significant findings. There were no clinically significant diagnoses identified from MRI in either group. While some incidental findings were identified, none were considered to be clinically significant and so no further follow up action was needed (see Table 1).

**Table 1:**
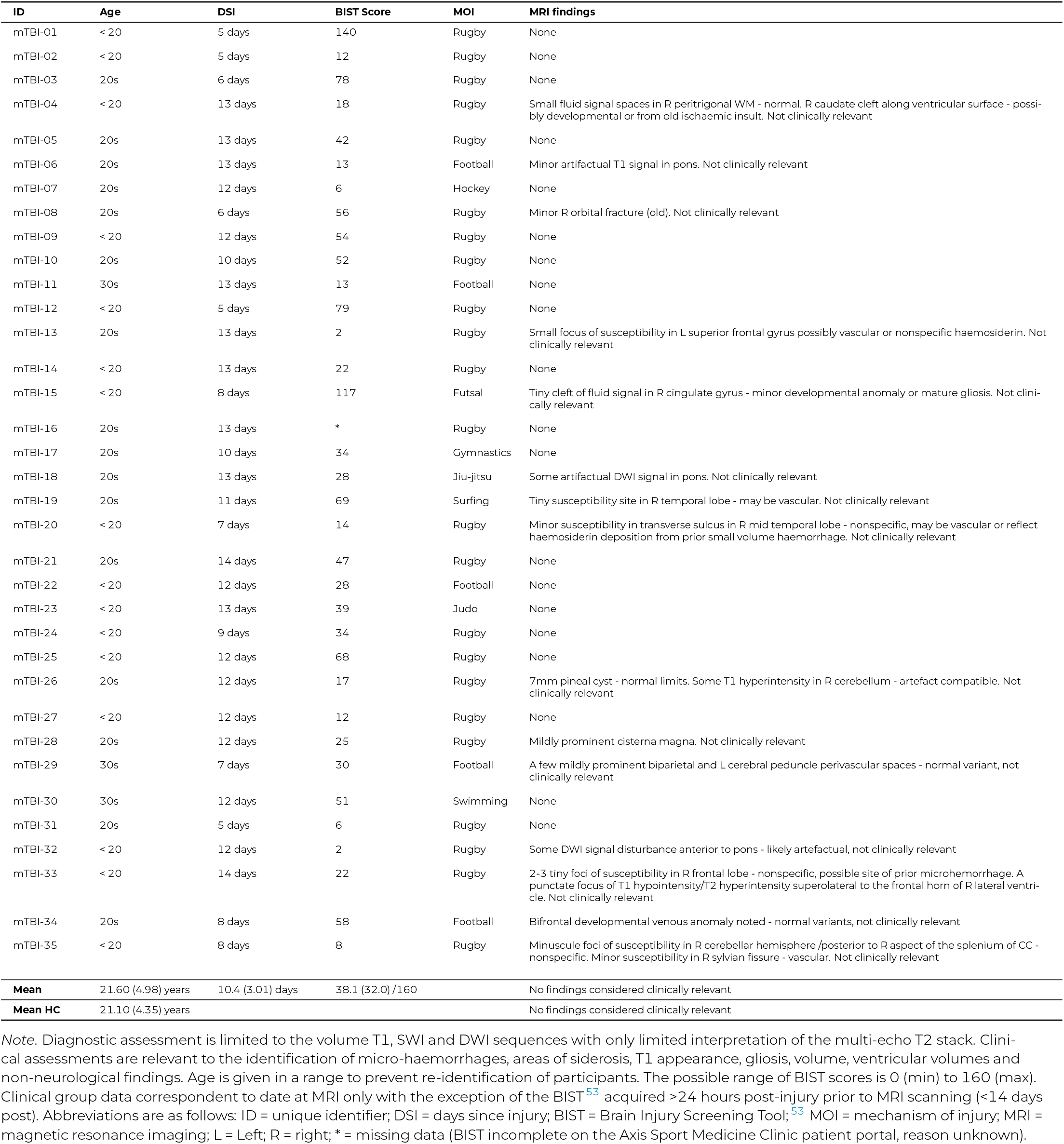
Summary of sr-mTBI participant clinical characteristics.

| ID | Age | DSI | BIST Score | MOI | MRI findings |
| --- | --- | --- | --- | --- | --- |
| mTBI-01 | < 20 | 5 days | 140 | Rugby | None |
| mTBI-02 | < 20 | 5 days | 12 | Rugby | None |
| mTBI-03 | 20s | 6 days | 78 | Rugby | None |
| mTBI-04 | < 20 | 13 days | 18 | Rugby | Small fluid signal spaces in R peritrigonal WM - normal. R caudate cleft along ventricular surface - possibly developmental or from old ischaemic insult. Not clinically relevant |
| mTBI-05 | 20s | 13 days | 42 | Rugby | None |
| mTBI-06 | 20s | 13 days | 13 | Football | Minor artifactual T1 signal in pons. Not clinically relevant |
| mTBI-07 | 20s | 12 days | 6 | Hockey | None |
| mTBI-08 | 20s | 6 days | 56 | Rugby | Minor R orbital fracture (old). Not clinically relevant |
| mTBI-09 | < 20 | 12 days | 54 | Rugby | None |
| mTBI-10 | 20s | 10 days | 52 | Rugby | None |
| mTBI-11 | 30s | 13 days | 13 | Football | None |
| mTBI-12 | < 20 | 5 days | 79 | Rugby | None |
| mTBI-13 | 20s | 13 days | 2 | Rugby | Small focus of susceptibility in L superior frontal gyrus possibly vascular or nonspecific haemosiderin. Not clinically relevant |
| mTBI-14 | < 20 | 13 days | 22 | Rugby | None |
| mTBI-15 | < 20 | 8 days | 117 | Futsal | Tiny cleft of fluid signal in R cingulate gyrus - minor developmental anomaly or mature gliosis. Not clinically relevant |
| mTBI-16 | 20s | 13 days | * | Rugby | None |
| mTBI-17 | 20s | 10 days | 34 | Gymnastics | None |
| mTBI-18 | 20s | 13 days | 28 | Jiu-jitsu | Some artifactual DWI signal in pons. Not clinically relevant |
| mTBI-19 | 20s | 11 days | 69 | Surfing | Tiny susceptibility site in R temporal lobe - may be vascular. Not clinically relevant |
| mTBI-20 | < 20 | 7 days | 14 | Rugby | Minor susceptibility in transverse sulcus in R mid temporal lobe - nonspecific, may be vascular or reflect haemosiderin deposition from prior small volume haemorrhage. Not clinically relevant |
| mTBI-21 | 20s | 14 days | 47 | Rugby | None |
| mTBI-22 | < 20 | 12 days | 28 | Football | None |
| mTBI-23 | < 20 | 13 days | 39 | Judo | None |
| mTBI-24 | < 20 | 9 days | 34 | Rugby | None |
| mTBI-25 | < 20 | 12 days | 68 | Rugby | None |
| mTBI-26 | 20s | 12 days | 17 | Rugby | 7mm pineal cyst - normal limits. Some T1 hyperintensity in R cerebellum - artefact compatible. Not clinically relevant |
| mTBI-27 | < 20 | 12 days | 12 | Rugby | None |
| mTBI-28 | 20s | 12 days | 25 | Rugby | Mildly prominent cisterna magna. Not clinically relevant |
| mTBI-29 | 30s | 7 days | 30 | Football | A few mildly prominent biparietal and L cerebral peduncle perivascular spaces - normal variant, not clinically relevant |
| mTBI-30 | 30s | 12 days | 51 | Swimming | None |
| mTBI-31 | 20s | 5 days | 6 | Rugby | None |
| mTBI-32 | < 20 | 12 days | 2 | Rugby | Some DWI signal disturbance anterior to pons - likely artefactual, not clinically relevant |
| mTBI-33 | < 20 | 14 days | 22 | Rugby | 2-3 tiny foci of susceptibility in R frontal lobe - nonspecific, possible site of prior microhemorrhage. A punctate focus of T1 hypointensity/T2 hyperintensity superolateral to the frontal horn of R lateral ventricle. Not clinically relevant |
| mTBI-34 | 20s | 8 days | 58 | Football | Bifrontal developmental venous anomaly noted - normal variants, not clinically relevant |
| mTBI-35 | < 20 | 8 days | 8 | Rugby | Minuscule foci of susceptibility in R cerebellar hemisphere /posterior to R aspect of the splenium of CC - nonspecific. Minor susceptibility in R sylvian fissure - vascular. Not clinically relevant |
| **Mean** | 21.60 (4.98) years | 10.4 (3.01) days | 38.1 (32.0) /160 |  | No findings considered clinically relevant |
| **Mean HC** | 21.10 (4.35) years |  |  |  | No findings considered clinically relevant |

Note. Diagnostic assessment is limited to the volume T1, SWI and DWI sequences with only limited interpretation of the multi-echo T2 stack. Clinical assessments are relevant to the identification of micro-haemorrhages, areas of siderosis, T1 appearance, gliosis, volume, ventricular volumes and non-neurological findings. Age is given in a range to prevent re-identification of participants. The possible range of BIST scores is 0 (min) to 160 (max). Clinical group data correspondent to date at MRI only with the exception of the BIST<sup>53</sup> acquired >24 hours post-injury prior to MRI scanning (<14 days post). Abbreviations are as follows: ID = unique identifier; DSI = days since injury; BIST = Brain Injury Screening Tool;<sup>53</sup> MOI = mechanism of injury; MRI = magnetic resonance imaging; L = Left; R = right; \* = missing data (BIST incomplete on the Axis Sport Medicine Clinic patient portal, reason unknown).

### Neuroimaging

Details on image acquisition and processing have been previously reported,^45^ and are summarised here for brevity.

### Acquisition

MRI data were acquired on a 3T Siemens MAGNETOM Vida Fit scanner (Siemens Healthcare, Erlangen, Germany) equipped with a 20-channel head coil. A 3D flow-compensated GRE sequence was used to obtain magnitude and unfiltered phase images suitable for QSM reconstruction. Data were collected at 1 mm isotropic voxel size with matrix size = 180 x 224 x 160 mm, TR = 30 ms; TE = 20 ms; FA = 15°; FoV = 224 mm in a total acquisition time of ~ 3.43 minutes. For each participant, a high-resolution 3D T1-weighted (T1w) anatomical image volume was acquired for coregistration, parcellation and segmentation using a Magnetisation-Prepared Rapid Acquisition Gradient Echo (MPRAGE) sequence (TR = 1940.0 ms; TE = 2.49 ms, FA= 9°; slice thickness = 9 mm; FoV = 230 mm; matrix size = 192 x 512 x 512 mm; GRAPPA = 2; voxel size 0.45 x 0.45 x 0.90 mm) for a total acquisition time of ~ 4.31 minutes. DICOMs were converted to NIfTI files and transformed to brain imaging data structure (BIDS)^54^ for further processing using *Dcm2Bids*^55^ version 3.1.1, which is a wrapper for *dcm2niix*^56^ (v1.0.20230411).

### Image processing

Bias field-corrected^57,58^ T1w images were processed in FreeSurfer^59^ to 1) delineate pial and grey matter/white matter (GM/WM) boundary meshes, and; 2) generate estimates of cortical thickness and curvature for each vertex.^60^ QSM images were reconstructed using a rapid open-source minimum spanning tree algorithm (ROMEO),^61^ background field removal with projection onto dipole fields (PDF),^**?**^ and sparsity-based rapid two-step dipole inversion (RTS);^62^ a pipeline congruent with recent consensus statement recommendations for best-practice QSM reconstruction.^63^ All QSM reconstruction was carried out via QSMxT^64^ v6.4.2 (https://qsmxt.github.io/QSMxT/) and used a robust two-pass combination method for hole-filling and artefact reduction.^65^

Subsequent processing was performed using FSL.^66–68^ For each subject, the raw magnitude image was skull-stripped^69^ and binarised. These binary masks were used to erode non-brain signal around the brain perimeter using *fslmaths*. Magnitude images were linearly coregistered to the T1w image using FMRIB’s Linear Transformation Tool (*FLIRT*)^70–72^ with 12 degrees of freedom. Due to variability in acquisition type, FoV and matrix size between subjects’ QSM and T1w images, the 12 DoF linear registration provided more accurate alignment compared to the 6 DoF alternative, allowing for better compensation of non-rigid anatomical variations upon visual inspection. The resulting transformation matrix was used for spatial normalisation of the QSM images to T1w space. In line with prior research,^60^ QSM maps were then thresholded into separate inter-voxel sign (positive and negative) maps with *fslmaths*. Traditional QSM maps already represent average voxel-wise susceptibility values,^73^ this averaging can obscure individual susceptibility sources as well as introduce further confounding effects when further averaging across voxels during analysis. Thresholding may help address the latter limitation by isolating voxels containing paramagnetic substrates, such as iron, which could enable more targeted analyses of these specific susceptibility sources. Only positive sign maps were used in further processing and analyses to target cortical iron distribution.

### Cortical column generation

To generate cortical columns and sample positive susceptibility values, we adapted a pipeline previously applied to DWI^74^ and QSM^45,60^ for depth- and curvature-specific cortical analysis. First, the T1w FreeSurfer^59^ recon served as an input into the *easy_lausanne* tool (https://github.com/mattcieslak/easy_lausanne.git) based on the open-source Connectome Mapper^75^ to separate the cortex into 34 ROIs per hemisphere according to the Lausanne multi-scale atlas (equivalent to the Desikan-Killiany atlas^76^ native to FreeSurfer).^59^

### Depth

Cortical columns were created for each hemisphere in T1w space with *write_mrtrix _tracks*^77^ in MATLAB (version R2024a), which was used to connect vertex pairs between the pial and GM/WM boundary surface meshes. Each cortical column was segmented into 6 equidistant depths extending from the pial surface to the GM/WM boundary using MRtrix3 *tckresample*.^77^ This approach differs from previous studies utilising this technique, where 21 equidistant depths were employed.^45,60,74^ Here, we used 6 depths to decrease the number of depth-wise comparisons, and to better approximate the structure of the intracortical layers.^78,79^ It should be noted here that these represent equidistant segmentations rather than specific cellular laminae (L1 to L6) of the cortex as it is important to distinguish results produced using this approach from ultra-high field investigations of cyto- and myelo-architecture in the cerebral cortex; results described herein are related to cortical *depth*, rather than *layer*.

### Curvature

The columns were also categorised based on cortical curvature, derived from FreeSurfer’s^59^ Gaussian curvature values at each GM/WM boundary vertex^80^ and quantified in units of 1/mm^2^. The categories included the gyral crown (curvature values: −0.6 to −0.1), sulcal bank (−0.1 to 0.1), sulcal fundus (0.1 to 0.6), or whole-ROI (−0.6 to 0.6).^60^ Positive curvature values indicated sulci, while negative values indicated gyri, with higher values corresponding to deeper curvatures.^60^ Only columns ranging from 0.5 mm to 6 mm in length were included in the analysis to capture plausible cortical morphology.^81^ Depth was measured in percentage of cortical thickness rather than absolute metrics (mm) to mitigate any variability between participants.

### Personalised QSM profiles

We generated individual QSM profiles for each ROI at the bilateral level using MATLAB (2024a) (see Figure 1 for visualisation). Mean positive susceptibility values were extracted across the whole ROI (curvature and depth combined), as well as 3 curvature bins (gyral crown, sulcal bank, and sulcal fundus) and 6 cortical depths independently for all 34 ROIs. For the whole-ROI profiles, z-scores were calculated for all participants (both healthy controls and mTBI), by subtracting the HC group mean from each individual’s susceptibility value and dividing by the HC group standard deviation; a method commonly used in prior research.^47,48,51,52^To bring the HC data closer to a normal distribution, outlier scores for the HC group were filtered^47^ if they fell outside 2 times the interquartile range (IQR), a more stringent criterion than the methods used to identify mild outliers 1.5 times the IQR, but less extreme than the more conservative filter of 3 times the IQR.^82^ As a result, data from *n = 1* HC participants were excluded in 3 of the 34 ROIs, and data from *n = 2* HC participants were excluded in 1 of the 34 ROIs. After filtering, the Shapiro-Wilk normality test yielded an average *W* value of *M* = 0.96 (*SD* = 0.02) across z-distributions for all ROIs, indicating that the data distribution within each ROI closely approximates normality. The final equation for deriving the whole-ROI z-scores for individual mTBI participants was as follows:

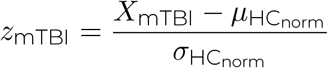

where *z*_mTBI_ represents the ROI-wise z-score for each individual mTBI participant; *X*_mTBI_ is the ROI-wise mean QSM value for each mTBI participant; *μ*_HCnorm_ is the mean ROI-wise QSM value of the HC group after outlier filtering to normalise the distribution, and; *σ*_HCnorm_ is the ROI-wise standard deviation of the HC group QSM values after outlier filtering. This approach ensures that mTBI participants’ susceptibility values are directly comparable to the healthy range reflecting a normalised distribution, allowing the detection of deviations that might reflect underlying pathology.

**Figure 1:**
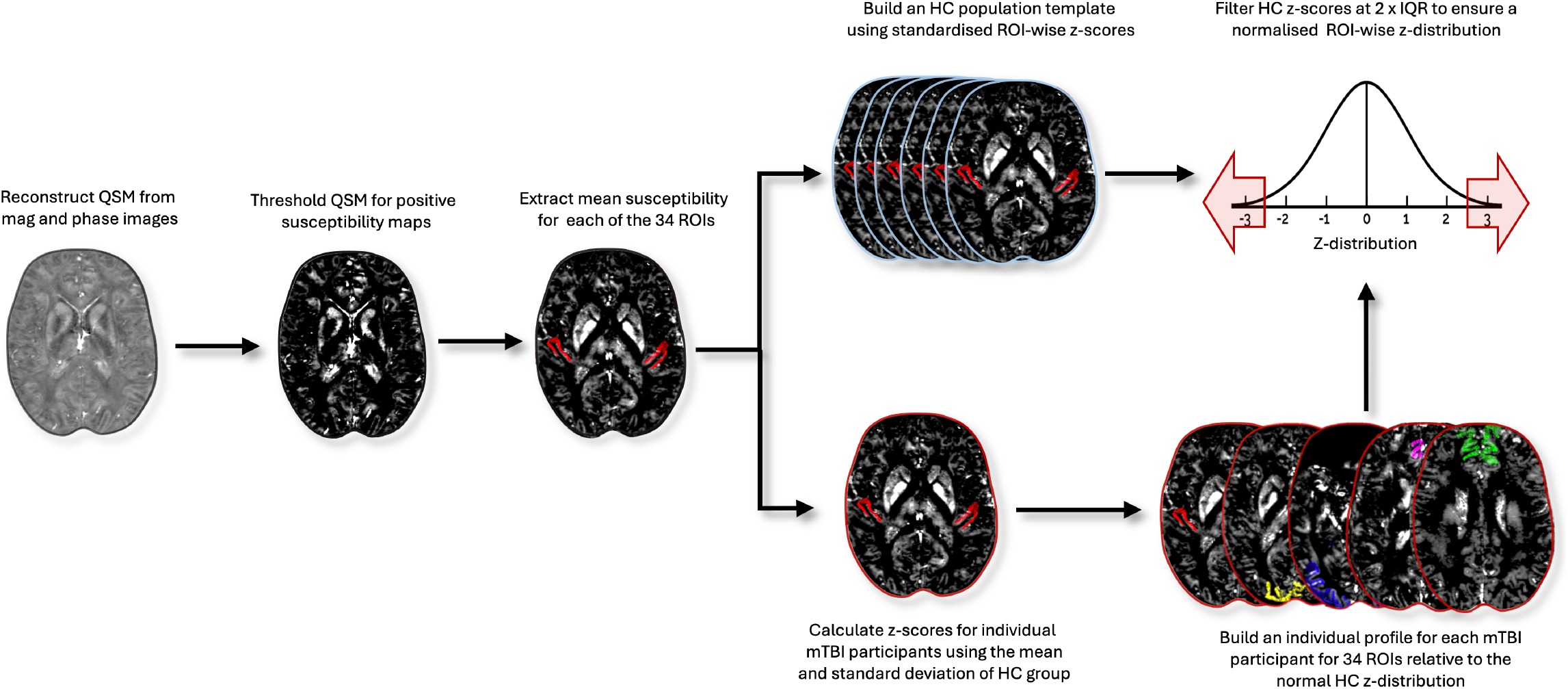
QSM post-processing and generation of individual iron profiles. Steps are performed after QSM image reconstruction using QSMxT.^65^ QSM images were thresholded to create a positive sign map, and mean susceptibility values were extracted for each ROI, as well as for each cortical depth (1 through 6) and curvature (crown, bank, and fundus). Z-scores were calculated using the mean and standard deviation of the healthy control group, and standardised around a mean of zero. The healthy control distribution was then filtered to remove outliers exceeding two times IQR, normalising the distribution. Individual profiles for mTBI participants were constructed by comparing each participant’s z-scores to the healthy normal distribution, while controlling for multiple comparisons across the 34 cortical ROIs. Abbreviations are as follows: QSM = quantitative susceptibility mapping; ROI = region of interest; HC = healthy control; IQR = interquartile range.

## STATISTICAL ANALYSES

To assess statistical significance for whole-ROI mTBI z-scores, two-tailed p-values were calculated from the z-scores using the cumulative distribution function of the standard normal distribution. A false discovery rate (FDR) correction 83 was applied to the p-values for 34 ROI-wise comparisons. The same process was repeated for each depth at each curvature bin, however, the IQR filter was omitted due to the number of comparison points.

To conduct secondary exploratory statistical tests, we divided the mTBI group into two subgroups: those whose z-scores significantly deviated from HC norms in the whole-ROI analysis (i.e., iron-abnormal) and those whose scores did not (i.e., iron-normal). Although there was no statistically significant difference in age between mTBI participants and controls, we performed a one-way analysis of variance (ANOVA) across these three groups to confirm that age was not driving the observed results at the individual level. Additionally, we used an independent samples t-test to assess whether injury severity (BIST^53^ scores) differed significantly between iron-abnormal and iron-normal mTBI participants, excluding mTBI-16 for this analysis only due to missing BIST data.

## RESULTS

### Regional individualised cortical iron profiles

We calculated personalised profiles of iron-related differences in positive susceptibility across 34 cortical ROIs for each mTBI participant, to understand the effects of mild brain trauma at the individual-level. Of the 35 mTBI participants, 11 (31%) exhibited significantly elevated positive susceptibility for at least one ROI relative to the healthy control population template (see Table 2), likely indicating elevated iron. No clinical participants’ z-scores were significantly lower than the healthy control population.

**Table 2:**
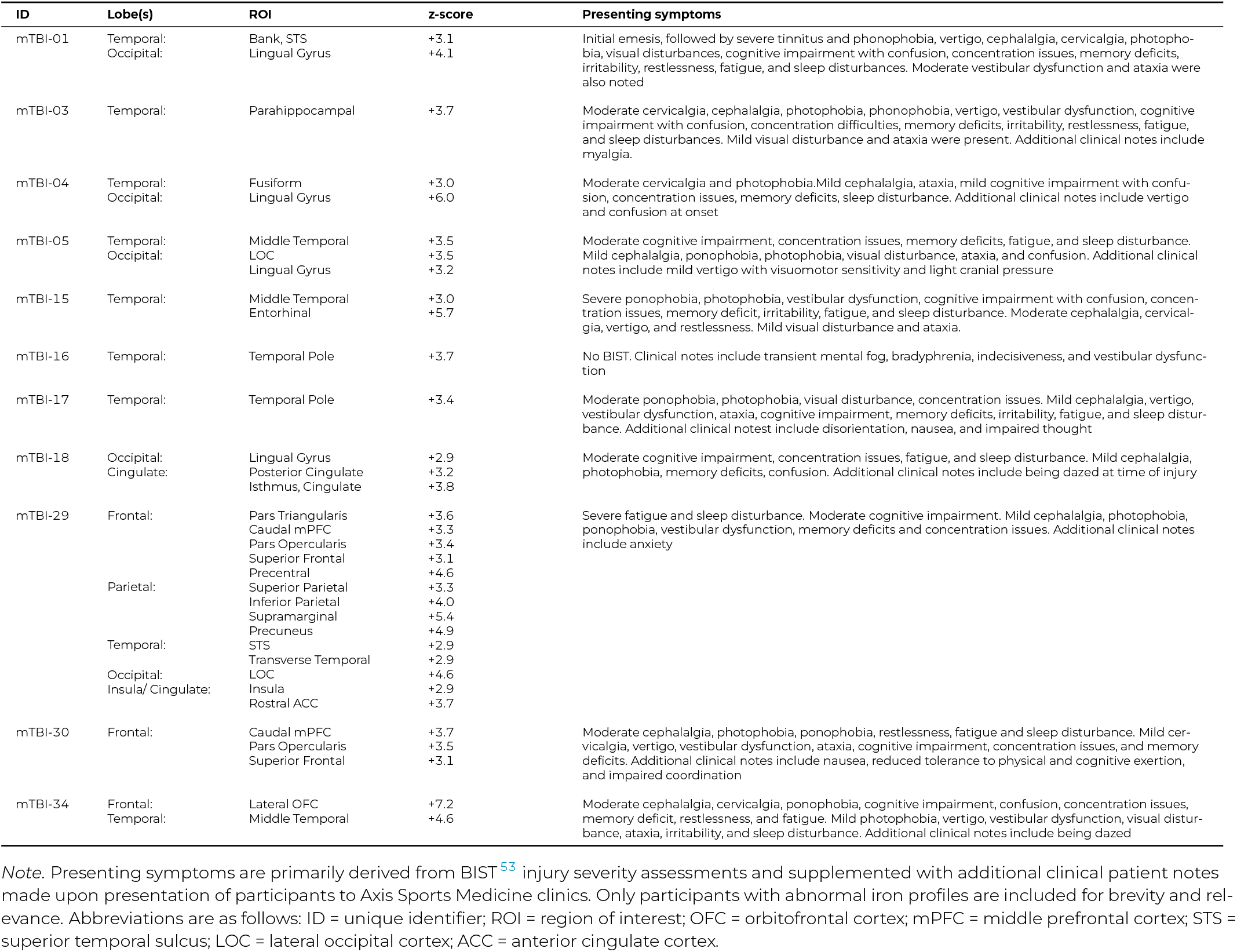
Summary of elevated ROI-wise susceptibility, z-score, and symptomatology for iron-abnormal sr-mTBI participants.

In these 11 clinical participants, injury-related elevated susceptibility was evident across all cortical lobes. Notably, a high density of affected ROIs was observed in the temporal lobe for 82% (9 out of 11) of participants with abnormal iron profiles (see Table 2). In contrast, 45% (5/11) had abnormal iron in occipital ROIs, 27% (3/11) in frontal ROIs, 18% (2/11) in the insula or cingulate, and only one participant (9%) had an abnormal profile inclusive of parietal ROIs. This variability reflects the diverse regions impacted by exposure to mild brain trauma (see Figure 2). For example, elevated cortical iron was localised to various temporal lobe ROIs in participants mTBI-03 (parahippocampal gyrus), mTBI-15 (middle temporal gyrus and entorhinal cortex), mTBI-16 (temporal pole), and mTBI-17 (temporal pole). In contrast, mTBI-30 showed extensive frontal lobe effects (caudal medial prefrontal cortex (mPFC), pars opercularis, and superior frontal gyrus), while the remaining participants exhibited multi-focal elevated positive susceptibility affecting two or more lobes (see Table 2 and Figure 2).

**Figure 2:**
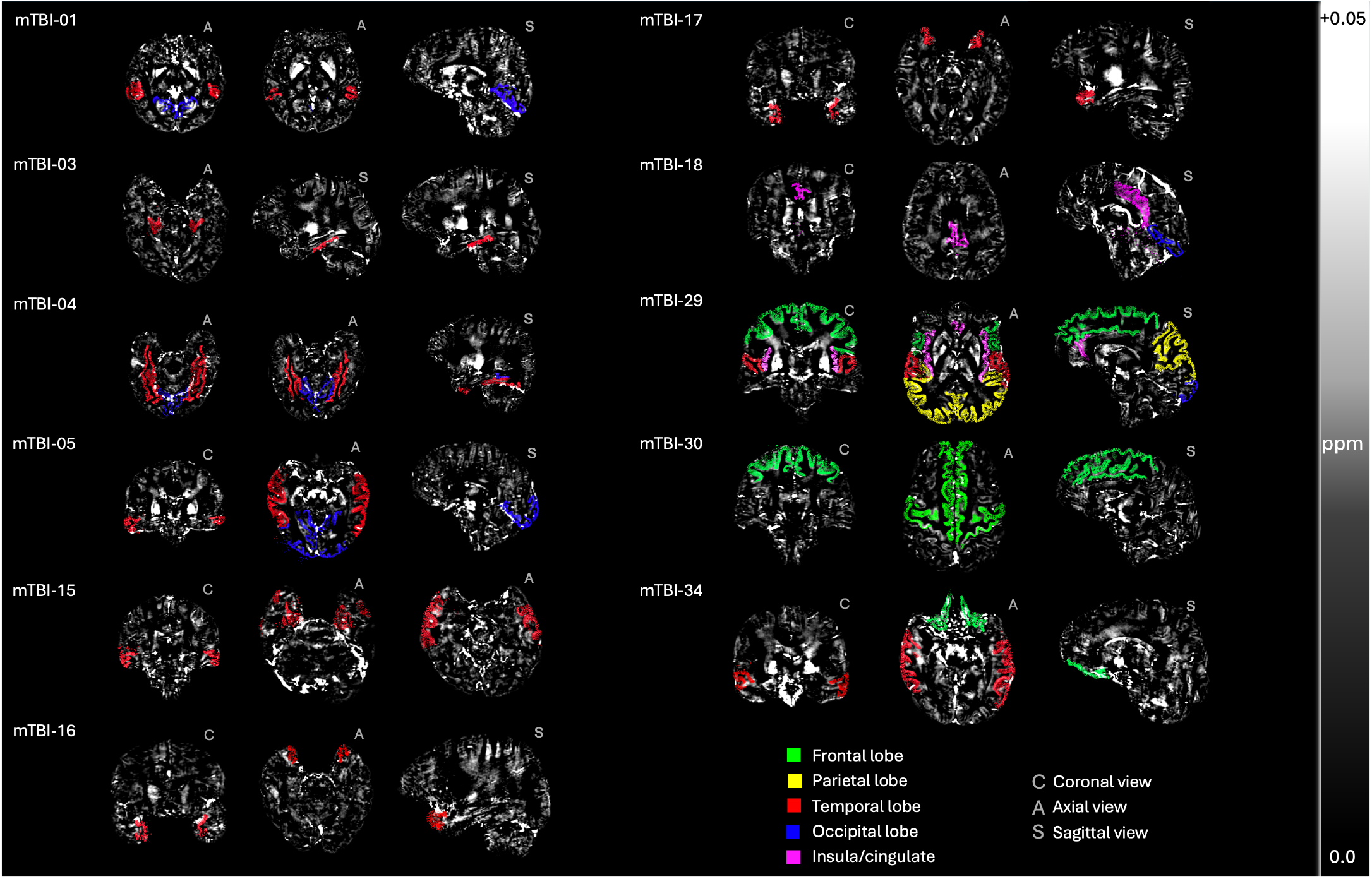
Individualised profiles of abnormal iron accumulation sites following mTBI. Visualisation of the specific ROIs and lobes affected in mTBI participants with significant deviations from the HC control group distribution, showing regions where z-scores significantly deviate from the healthy control population, highlighting the individualised profiles of iron accumulation following mTBI. The selected orientations (C: coronal, A: axial, S: sagittal) are for best visualisation of each participant’s result. These maps have been threshold for positive susceptibility values (iron-related) and are expressed in parts per million (ppm) from 0.0 to +0.05.

After subdividing the mTBI participants into iron-normal (24/35; 69%) and iron-abnormal (11/35; 31%) based on their individual ROI-wise profiles, we used a one-way ANOVA to test if age was significantly different between the three groups (iron-normal mTBI, iron-abnormal mTBI, and controls) as well as an independent samples t-test to understand whether injury severity (BIST^53^) was significantly different for iron-normal and iron-abnormal mTBI participants. The ANOVA yielded a non-significant effect of age, *F*(2, 26) = 2.00, *p* = .16. BIST^53^ scores were significantly higher for the iron-abnormal mTBI group (*M* = 59.6, *SD* = 40.5) than the iron-normal mTBI group (*M* = 29.2, *SD* = 23.3), *t*(32) = 2.77, *p* = < .01, suggesting a link between injury severity and elevated regional cortical iron deposition (see Figure 3).

**Figure 3:**
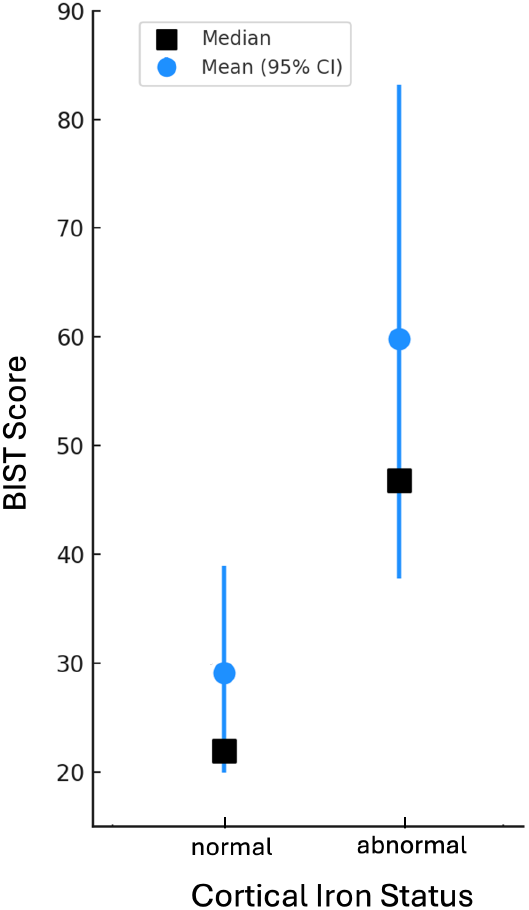
BIST scores by cortical iron status. *Note:* Brain Injury Screening Tool (BIST)^53^ scores as a function of cortical iron status (normal vs. abnormal) in mTBI participants. Median (black squares) and mean (blue circles) BIST^53^ scores were both higher for iron-abnormal mTBI participants. 95% confidence intervals are indicated using blue bars for each group.

### Exploratory depth- and curvature-specific individualised iron profiles

Exploratory analyses sensitive to cortical depth (1 through 6, from the pial surface to the GM/WM interface) and curvature (crown, bank, and fundus) further highlighted the heterogeneity of iron deposition in mTBI. Only 17% (6/35) of participants retained normal iron profiles; the remaining 83% (29/35) showed elevated susceptibility in at least one ROI, for at least one depth, and for at least one curvature. Isolated instances of negative z-scores (indicating lower iron compared to HC) were observed in 7 of the 29 participants, but were typically limited to a single ROI/depth combination. Overall, abnormal iron accumulation was most pronounced in the sulcal fundus, followed by the sulcal bank, and was least evident in the gyral crown (see Figure 4 for reference). These distributions were consistent across both superficial (1-2), mid (3-4), and deeper (5-6) cortical depths, however, each unique combination of depth and curvature for each participant reflects individual variability in iron deposition across the cortex. For example, abnormal iron deposition for mTBI-25 was exclusive to superficial depths (1 and 2) in the gyral crown and sulcal fundus of the iron-elevated ROIs. In contrast, mTBI-34 exhibited more widespread iron accumulation which became more pronounced at deeper depths, and was observed in the gyral crown, sulcal bank, sulcal fundus, or a combination thereof, depending on the ROI. Participant mTBI-29 showed iron accumulation at all depths but this was most pronounced at mid-to-deep depths (3-5) in the sulcal bank and fundus (see Figure 4).

**Figure 4:**
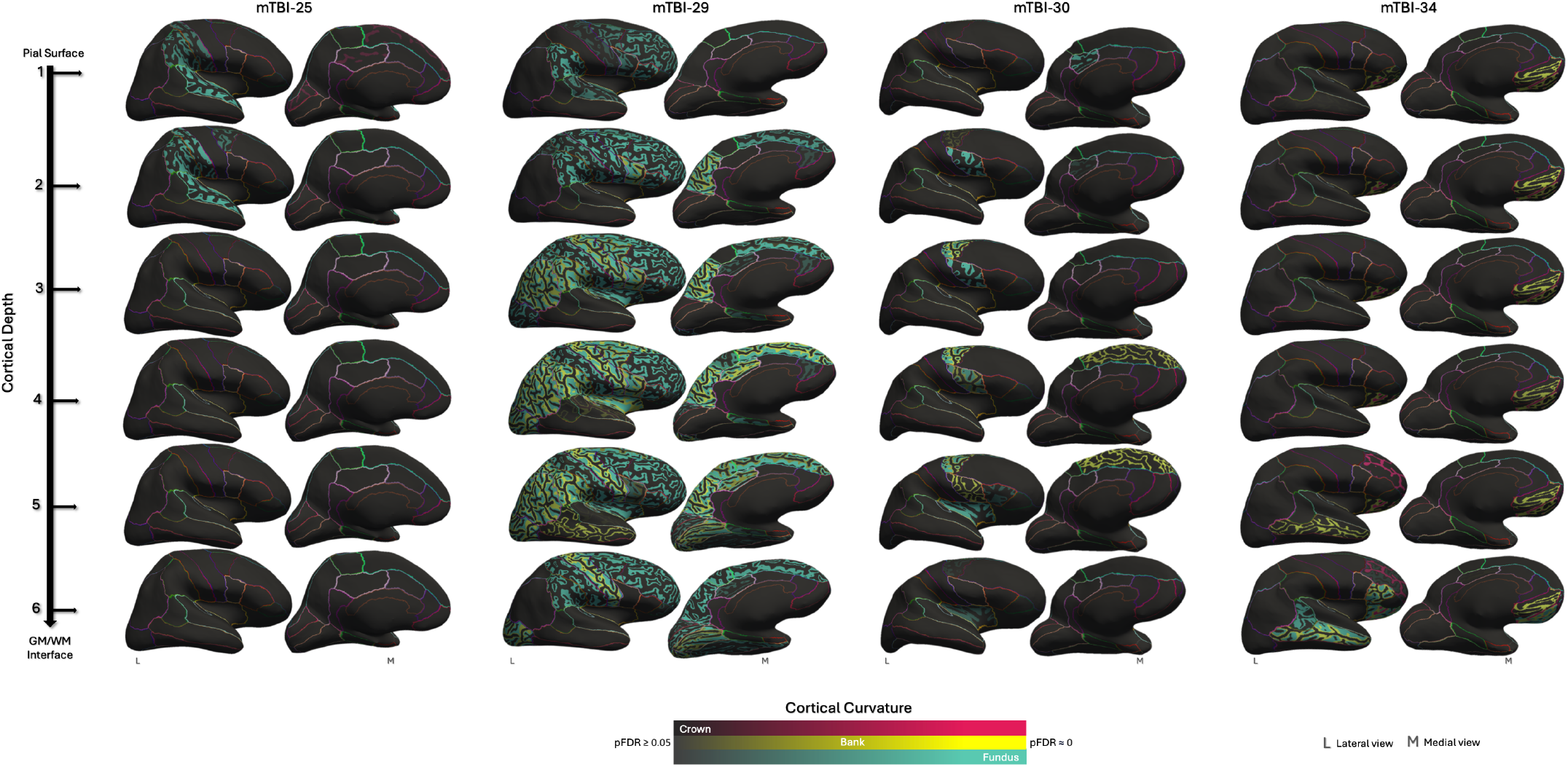
Cortical depth- and curvature-specific profiles of mTBI-related abnormal iron accumulation. Cortical depth- and curvature-specific iron profiles across five representative sr-mTBI participants. Inflated surfaces show each of the six cortical depths, from the pial surface (depth 1) to the GM/WM interface (depth 6). Regions of abnormal iron deposition are colour-coded according to cortical curvature (crown = pink, bank = yellow, and fundus = blue). Colour intensity shows level of statistical significance (pFDR < 0.05). Lateral (L) and medial (M) views are used to visualise the whole brain. Boundaries between ROIs are delineated using coloured lines.

For the 11 participants who demonstrated increased iron in the initial whole-ROI analyses, a similar ROI-wise distribution of elevated iron was observed in the depth- and curvature-specific analyses. For example, in whole-ROI analyses mTBI-30 showed elevated susceptibility in the caudal mPFC, pars opercularis, and superior frontal gyrus (see Table 2 and Figure 2). Depth and curvature analyses revealed that iron deposition in the caudal mPFC occurred across superficial-to-deep depths (2-5) in the sulcal bank and at mid-to-deep depths (3-6) in the sulcal fundus, with no deviation from the control template at the gyral crown (see Figure 4). In the pars opercularis, iron accumulation was apparent at mid-to-deeper depths (4-5) in the bank and at superficial-to-mid depths (2-4) in the sulcal fundus, again with no accumulation at the gyral crown. Similarly, the superior frontal region showed mid-to-deep (4-5) abnormalities in the bank. Regions that did not appear in the initial analyses, including the insula, lateral occipital cortex (LOC), pars triangularis, posterior cingulate, precentral, and superior parietal areas, exhibited elevated iron in depth- and curvature-specific analyses, mostly concentrated in the sulcal bank or fundus. Conversely, mTBI-25 exemplifies a case where no abnormal iron profile was detected in the whole-ROI analysis, but became evident with analyses sensitive to anatomical morphology (see Figure 4).

## DISCUSSION

Previous studies using QSM to examine the role of brain iron following mTBI have primarily focused on subcortical brain areas or global grey and/or white matter.^36–44^ Only two investigations have included cortical ROIs and, of these, only our previous work^45^ accounted for anatomical variations in cortical depth and curvature. However, these studies relied on group-level statistical analyses, which can obscure individual brain changes due to the subtle nature of cell damage associated with mTBI. This approach may limit our understanding of this heterogeneous injury and inhibits the identification of personalised injury profiles, hindering the implementation of individualised rehabilitation strategies and treatments. We believe, therefore, that research that incorporates comparisons of individual clinical participant data to healthy normative ranges can play a key role in informing individualised neural and pharmacological interventions. We conducted the first investigation of individual differences in cortical magnetic susceptibility after sports-related mTBI across 34 cortical ROIs, using a healthy population template as a reference. Secondary exploratory analyses of individual differences in susceptibility distribution at different cortical depths and curvatures were included to better characterise injury profiles for each participant

Our findings revealed that a substantial subset of individuals with mTBI exhibit elevated levels of cortical magnetic susceptibility, indicating injury-related iron accumulation. Our primary investigation evidenced abnormal iron profiles in just under one-third (31%; 11/35) of participants relative to the healthy control population. In these 11 ‘iron-abnormal’ clinical participants, elevated susceptibility was evident across all cortical lobes, with a notably higher density of affected ROIs localised to the temporal lobe. At the individual level, elevated iron profiles were either focal to specific lobes (such as the temporal lobe in mTBIs −03, −15, −16, and −17, and widespread frontal anomalies in mTBI-30) or bi-focal (affecting two lobes). Only one participant (mTBI-29) exhibited multi-focal cortex-wide elevated susceptibility involving the frontal, temporal, parietal, occipital, and insular/cingulate cortices. These results suggest that whilst there is significant inter-subject variability, iron accumulation after mTBI preferentially affects temporal regions. Additionally, we found that mTBI participants with abnormal iron accumulation experienced more severe symptoms. Taken together, our findings support an iron-related mechanism of secondary injury that modulates symptom presentation.

Iron dyshomeostasis in the basal nuclei is known to impair cognitive function after mTBI,^16,17^ however, little is known about the effect of *cortical* iron aggregation on mTBI symptomatology. In our previous work,^45^ we reported few correlations between regions of cortical iron accumulation and BIST^53^ scores, a measure of injury severity and dominant symptom cluster (physical, vestibular, and cognitive). This lack of correlation may be accounted for by known variability in the accuracy of symptom reporting or purposeful underreporting of symptoms, a phenomenon notorious among sports players.^84^ However, group-level examinations may also obscure individual differences in addition to inhibiting the implementation of more appropriate statistical approaches. By assessing the effect of mTBI at the individual level, we were able to facilitate more precise between-group analyses of injury severity by differentiating between iron-normal and iron-abnormal mTBI participants. Our results revealed a significantly higher symptom burden for participants with mTBI when their iron profiles were also abnormal.

The field generally lacks reliable correlations between subjective assessments of injury severity and objective measures of brain injury and recovery,^85^ as well as alignment between cognitive or clinical findings and neuroimaging results.^86^ Identifying reliable, objective markers of structural changes that are related to subjective self-reported symptoms is crucial because individual variations in brain injury location and severity can lead to vastly different clinical presentations and recovery trajectories but may be missed in group-level analyses.^8^ Although research indicates that most individuals recover well from mTBI, between 15%^11^ and 30%^8^ of patients experience significant, and in some cases life-changing, long-term clinical sequelae. Understanding the underlying pathophysiological drivers of these poorer outcomes is essential for enabling precise, patient-specific clinical interventions. We found that 31% of participants exhibited abnormal iron profiles substantial enough to be detected in the primary ROI-wise analyses, which were linked to poorer outcomes. This aligns with findings from other individualised studies, which have reported that a similar percentage (28%) of participants with ms-TBI exhibit white matter anomalies in the subacute phase, associated with worse cognitive outcomes than participants without.^51^ In addition, standardised susceptibility values in the basal nuclei are reported to correlate with mTBI symptom duration, but only in a sub-group of participants with persistent symptomatology for at least a week.^40^ In keeping with these findings, our results reinforce the importance of individualised analyses in revealing associations between the extent of microstructural pathology and negative outcomes following mTBI as well as the ability of this approach to differentiate at-risk sub-cohorts. Here, the value of personalised approaches to understanding mild brain injury becomes strikingly apparent. Further longitudinal studies that track participants through recovery could help to determine whether elevated iron levels are associated with prolonged recovery, persistent post-concussive symptoms, or adverse outcomes later in life. Such research may offer insights into why a subset of individuals with mTBI fail to recover fully.

The promise of individualised assessments to identify biomarkers for mild brain injury is particularly salient given the current absence of objective markers for mTBI diagnosis. Diagnostic decisions are limited to subjective self-report and assessments of physiological function,^13,85^ as heterogeneity of mTBI complicates efforts to identify reliable biomarkers or imaging signatures that can be applied universally across patients. Whether cortical iron accumulation reflects inflammatory processes, BBB disruption,^9^ ferroptosis,^31,87^ or other cytotoxic processes is beyond the scope of the current research. However, we posit that the significantly higher symptom burden observed in the iron-abnormal mTBI cohort supports an iron-mediated mechanism of brain changes related to injury severity and functional impairment, and marks iron as a promising mTBI biomarker. Of particular note, many of the symptoms observed in mTBI resemble those seen in other iron-mediated processes, such as cognitive decline in normal aging,^88^ and the cognitive and motor dysfunctions characteristic of neurodegenerative diseases like Alzheimer’s, Huntington’s, Parkinson’s, and Friedrich’s ataxia, as well as multiple sclerosis, where iron dysregulation is a hallmark feature.^9,89^ Genetic disorders of iron over-load, such as neuroferritinopathy, also present with cognitive and motor symptoms.^90^

The susceptibility of the temporal lobe to iron accumulation post-mTBI, evident in this and previous research,^45^ aligns with the memory impairments characteristic of mTBI.^91^ Individual-level data from our mTBI sample emphasises the link between high temporal iron accumulation and memory deficits, as exemplified by BIST^53^ scores related to memory (i.e., mTBIs −01, −03, −04, −05, −15, −17, −29, and −34; see Table 2). The distributions of p-tau focal to temporal (and frontal) cortex are considered pathognomonic of CTE,^92,93^ an mTBI-related neurodegenerative disorder. Co-localisation of iron with p-tau in CTE has been highlighted in histological examinations.^34^ Whilst the precise relationship between iron overload and the downstream hyperphosphorylation of tau proteins remains an active area of research, further exploration of the interplay between acute cortical iron elevation, symptom burden, temporal recovery dynamics, and long-term brain health outcomes is warranted.

The diverse range of mechanistic antecedents, pathological mechanisms, and clinical outcomes associated with mTBI reflects the complexity of the underlying pathophysiology.^8^ mTBI is not a uniform injury, and structural indicators, such as those commonly observed in other forms of TBI, do not always correlate with clinical symptoms or outcomes.^94^ Here, we speculate that abnormal iron accumulation in specific cortical ROIs may be related to participant symptomatology. For instance, the superior temporal sulcus (STS) plays a key role in social cognition, empathy, mentalising about others’ emotional states, and ‘theory of mind.’^95–97^ Structural changes to this region may explain the severe irritability reported by mTBI-01 (see Table 2), along with complaints of severe tinnitus.^98^ As a hub for audiovisual integration,^99^ the superior temporal region could also be involved in phonophobia (sound sensitivity), visual disturbances, and vestibular dysfunction;^100^ symptoms experienced by both mTBI-01 and mTBI-29 whose profiles indicated local elevations in cortical iron. Similarly, the lingual gyrus is active during migraine episodes and responds to luminous stimuli, suggesting its involvement in photophobia, visual processing anomalies, and cephalalgia (headache).^101^ These symptoms were observed in all clinical participants (mTBIs −01, −04, −05, −18; see Table 2) with iron aggregation in this region. Our findings allude to specific areas of cortical iron accumulation that show a relationship to clinical sequelae, suggesting that regions with higher iron burden may be evidence of microstructural cell damage that disrupts normal function. Future research should integrate functional MRI to enhance the translation of findings and improve the mapping of structural changes to deficits in functional connectivity and brain networks. Without additional research, observations about regions of iron accumulation and symptom presentation are speculative and much less convincing than case-matching between brain lesions in gross TBI and neurobehavioural symptom presentation.^102^

### Depth- and curvature-specific iron accumulation

Secondary analyses revealed significant inter-individual heterogeneity in depth- and curvature-specific cortical iron accumulation, however, there was a consistent trend for iron deposition in the sulci, particularly concentrated at the fundus, followed by the sulcal bank. This pattern may be attributed to the heightened vulnerability of the sulcal fundus to mTBI-related injury, which is susceptible to mechanical deformation due to the ‘water hammer effect,’ where cerebrospinal fluid (CSF) is forced into the depths of the sulci, causing local damage.^103^ Supporting this, previous research has demonstrated that mTBI increases cortical curvature in the sulcus,^104^ as well as widens the sulci and causes focal vascular injury and microhemorrhages in the fundus, as evidenced by susceptibility-weighted imaging (SWI).^103^ Conversely, in more severe TBI, contusions are often concentrated at the gyri,^91^ suggesting that gyral iron accumulation may represent a less severe version of this type of injury. This highlights how crucial injury biomechanics may be to understanding antecedents to differences in the location of neuropathology. Research shows that different types of head impacts can result in varying brain deformations and injury patterns, with sulci being particularly vulnerable to mechanical strain, which is consistent with, and can be predicted by, patterns of tauopathy observed in neurodegenerative conditions.^105^ Personalised iron accumulation patterns may provide insight into the specific injury mechanisms and related cellular disruption experienced by each individual participant at the acute stage. Elucidating this link, as well as how this may be related to mTBI-induced neuropathology in later life, should be a focus for future research.

Histological studies, including Perls iron staining and ultra-high-field (7T) R_2_* mapping of tissue samples^106^ have localised iron deposition to specific cortical layers, reflecting distinct cyto- and myelo-architectural features with layer-specific distributions that show congruence with *in-vivo* QSM.^107^ In healthy populations, iron concentrations typically increase from the pial surface to-ward the GM/WM boundary; deviations away from baselines for each layer suggest an injury-specific model of cortical cellular trauma. For instance, Layer I primarily contains axons and dendrites, with the cell bodies of these processes residing in deeper layers,^108^ and iron accumulation in different layers may point to diverse pathologies affecting different parts of the cell. Depth-wise patterns may also be related to injury biomechanics: superficial iron accumulation may be a result of perivascular trauma, which is often linked to microhemorrhages and microglial activation after mTBI,^27,28,91^ whereas deeper iron deposition may reflect more severe shear forces, which are known to cause significant damage near the GM/WM interface in mTBI.^109^ This is supported by computational modelling showing shear forces are concentrated in this region,^105^ which are also a common site of microbleeds.^110^ Tying this in with cortical curvature, this would most likely be apparent at the depths of the sulci.^103,105,111^ However, contusions of the cerebral gyri damage often follow a layer-specific pattern where most damage occurs at the superficial crest but extends through the cortical mantle to the GM/WM boundary in a ‘wedge’ of haemorrhage and necrotic tissue.^91^ Iron-dependent cell death, ferroptosis,^31,87^ may account for some of the similarities observed between this and less severe forms of TBI.

It is important to note that both injuries at the pial surface^111^ and closer to the GM/WM border^109^ have been related to adverse outcomes after mTBI. However, subpial iron deposition near small blood vessels in the fundus is congruent with the pathognomonic distributions of sulcal tauopathy seen in CTE,^111^ which may be related to breaches in the BBB.^105^ Injury-induced microvascular dysfunction may increase BBB permeability^112,113^ which likely increases active transport of non-heme iron via vascular endothelial cells into the superficial layers of the cortex^9,29,114^which could account for perivascular accumulations.^91,92^ The potential long-term effects of iron deposition in this young cohort warrants further investigation, given the known relationship between iron accumulation and tau pathology.^34^

## LIMITATIONS AND FUTURE RESEARCH

The ability of neuroimaging modalities to infer underlying biological processes is inherently limited, primarily by spatial constraints and inference about tissue composition from indirect markers. While the use of QSM can provide insight into potential iron accumulation, it remains a surrogate measure of iron-tissue content. Integrating complementary modalities, such as positron emission tomography (PET), would offer a more comprehensive understanding of the underlying neurobiology, for example by using tracers sensitive to inflammation or tau^115^ which may co-localise with iron overload. Additionally, the inclusion of blood-based biomarkers or other protein assays would enhance the interpretation of the neurobiological consequences of sr-mTBI, providing a clearer link between observed imaging abnormalities and molecular pathology. The cross-sectional nature of this study restricts our ability to establish a causal relationship between iron accumulation and symptom progression; here, longitudinal studies that track participants over time would be more effective in determining how iron deposition influences recovery and long-term outcomes, potentially also offering insights into the development of neurodegenerative conditions. This study does not address the precise mechanisms by which different types of mechanical strain or impact contribute to the observed depth- and curvature-specific patterns of iron accumulation. Whether these patterns are driven by depth-wise shear forces near the GM/WM interface^105,109^ or reflect superficial perivascular damage,^27,28,91^ or whether curvature-related deposition may be due to the ‘water hammer effect’ at the sulcal fundus^103^ or contusions of the gyri,^91^ or a combination of all of these factors, remains unclear. Future research should track athletes throughout the sporting season to better understand the specific biomechanics of their injuries at the time of insult. Longitudinal assessments here would also be useful for elucidating how patterns of iron deposition and the biomechanics of injury relate to the development of neuropathology across different cortical depths and layers.

It should be noted that this study did not control several potential confounding factors, including prior injuries, genetic predispositions, and environmental influences, all known to affect injury severity and recovery trajectories.^8^ Future studies may benefit from incorporating measures to account for these variables. Another limitation relates to the generation of a normal healthy control z-distribution, as there is no widely established best-practice method for this process. While this study filtered for outliers using robust techniques at two times the interquartile range, other individualised QSM research has employed less stringent criteria, such as filtering at three times the interquartile range to detect more severe TBI pathology.^47^ Future research would benefit from standardised practices for outlier detection. In addition, this research focused solely on net paramagnetic susceptibility voxel values, leaving net diamagnetic voxels, containing substances such as certain proteins or calcifications, unexplored. Previous studies in both mTBI^45^ and Alzheimer’s disease (AD)^60^ have suggested interactions between iron deposition and changes in diamagnetism on QSM. Future research should incorporate an analysis of both paramagnetic and diamagnetic susceptibility distributions to better characterise tissue changes after injury.

The current study also relied on a single-echo QSM sequence, which constrains susceptibility thresholding to a *between* voxels approach. Multi-echo sequences, which enable the separation of susceptibility sources *within* voxels,^107,116,117^ should be used in future research to further refine magnetic source separation. This approach would allow for more precise mapping of microstructural changes and improve the biological interpretation of the data. While this study approximated cortical architectonics by analysing six cortical depths, voxel resolution at a 3T field strength is limited and cannot delineate cortical laminae. Higher-field imaging (e.g., at 7T) would enable better resolution and alignment with cortical architectonics and should be considered for future research using a depth-wise approach to iron mapping. Finally, the focus on the male participants for this research limits the generalisability of the findings to females for whom hormonal variations,^118^ the use of oral contraceptives,^119^ and differences in neck musculature^120^ lend themselves to further heterogeneity in injury severity and outcome. Planned future research will extend these investigations to include female athletes to address this gap.

## CONCLUSIONS

To investigate the mechanisms of iron dyshomeostasis in acute mild traumatic brain injury, we conducted the first QSM study to assess individual profiles of cortical iron deposition. Our findings revealed significant heterogeneity in iron accumulation, which may be influenced by cortical morphology, highlighting the importance of examining mTBI at an individual level rather than relying on group analyses. This variability likely complicates the search for universal biomarkers, further underscoring the need for a personalised approach that integrates advanced imaging and detailed symptom profiling. Our results emphasise the need for more targeted, individualised interventions to improve outcomes based on personal profiling, and suggest that iron-mediated cell damage plays a key role in mTBI pathology.

## Data Availability

All data produced in the present study are available upon reasonable request to the authors

## AUTHOR CONTRIBUTIONS

**Christi A. Essex** (Conceptualization, Methodology, Project Administration, Validation, Software, Formal Analysis, Investigation, Resources, Data Curation, Writing - Original Draft, Writing - Review & Editing, Visualization); **Devon K. Overson** (Methodology, Software, Visualization, Validation, Writing - Review & Editing); **Jenna L. Merenstein** (Methodology, Writing - Review & Editing, Visualization); **Trong-Kha Truong** (Methodology, Software, Writing - Review & Editing); **David J. Madden** (Methodology, Software, Writing - Review & Editing); **Mayan J. Bedggood** (Writing - Review & Editing, Project administration, Investigation); **Catherine Morgan** (Methodology, Writing - Review & Editing); **Helen Murray** (Writing - Review & Editing); **Samantha J. Holdsworth** (Writing - Review & Editing); **Ashley W. Stewart** (Writing - Review & Editing); **Richard L.M. Faull** (Writing - Review & Editing); **Patria Hume** (Writing - Review & Editing); **Alice Theadom** (Conceptualization, Methodology, Writing - Review & Editing, Funding acquisition, Supervision); **Mangor Pedersen** (Conceptualization, Methodology, Writing - Review & Editing, Funding acquisition, Supervision)

## ACKNOWLEDGMENTS

We extend thanks to Amabelle Voice-Powell and Cassandra McGregor for their contribution to the data collection, and Tania Ka’ai for bringing her perspective to cultural considerations on this study. In addition, we thank Axis Sports Concussion Clinics, particularly Dr Stephen Kara, for their assistance with recruiting sr-mTBI participants and personnel at the Centre for Advanced Magnetic Resonance Imaging (CAMRI) for their assistance collecting MRI data. We also acknowledge Dr Tim Elliot for radiological reporting of all participants and Siemens Healthineers for the use of a work-in-progress (WIP) prototype sequence for the acquisition data used to perform QSM.

## DATA AVAILABILITY

De-identified MRI data and code used for image processing and statistical analysis can be made available upon request to the corresponding author. Parent codes for cortical column generation can be made available upon request from co-authors (JLM, TKT) based at the Brain Imaging and Analysis Center at Duke University Medical Center.

## REFERENCES

[1] Andrew I.R. Maas, David K. Menon, Geoffrey T. Manley, Mathew Abrams, Cecilia Åkerlund, Nada Andelic, Marcel Aries, Tom Bashford, Michael J. Bell, Yelena G. Bodien, Benjamin L. Brett, András Büki, Randall M. Chesnut, Giuseppe Citerio, David Clark, Betony Clasby, Jamie D. Cooper, Endre Czeiter, Marek Czosnyka, (..), and Roger Zemek. Traumatic brain injury: progress and challenges in prevention, clinical care, and research. The Lancet Neurology, 21(11):1004–1060, 11 2022.

[2] Emilie Isager Howe, Nada Andelic, Silje C.R. Fure, Cecilie Røe, Helene L. Søberg, Torgeir Hellstrøm, Øystein Spjelkavik, Heidi Enehaug, Juan Lu, Helene Ugelstad, Marianne Løvstad, and Eline Aas. Cost-effectiveness analysis of combined cognitive and vocational rehabilitation in patients with mild-to-moderate TBI: results from a randomized controlled trial. BMC Health Services Research, 22(1), 12 2022.

[3] Kevin M. Guskiewicz, Stephen W. Marshall, Julian Bailes, Michael Mccrea, Herndon P. Harding, Amy Matthews, Johna Register Mihalik, and Robert C. Cantu. Recurrent concussion and risk of depression in retired professional football players. Medicine and Science in Sports and Exercise, 39(6):903–909, 2007.

[4] Kerry McInnes, Christopher L. Friesen, Diane E. MacKenzie, David A. Westwood, and Shaun G. Boe. Mild Traumatic Brain Injury (mTBI) and chronic cognitive impairment: A scoping review. PLoS ONE, 12(4), 4 2017.

[5] Daniel F. Mackay, Emma R. Russell, Katy Stewart, John A. MacLean, Jill P. Pell, and William Stewart. Neurodegenerative Disease Mortality among Former Professional Soccer Players. New England Journal of Medicine, 381(19):1801–1808, 2019.

[6] Louis De Beaumont, Hugo Thoret, David Mongeon, Julie Messier, Suzanne Leclerc, Sebastien Tremblay, Dave Ellemberg, and Maryse Lassonde. Brain function decline in healthy retired athletes who sustained their last sports concussion in early adulthood. Brain, 132(3):695–708, 2009.

[7] Kevin M. Guskiewicz, Stephen W. Marshall, Julian Bailes, Michael McCrea, Robert C. Cantu, Christopher Randolph, and Barry D. Jordan. Association between Recurrent Concussion and Late-Life Cognitive Impairment in Retired Professional Football Players. Neurosurgery, 57(4):719–726, 2005.

[8] Sara B. Rosenbaum and Michael L. Lipton. Embracing chaos: The scope and importance of clinical and pathological heterogeneity in mTBI. Brain Imaging and Behavior, 6(2):255–282, 2012.

[9] Aleksandra Gozt, Sarah Hellewell, Phillip G.D. Ward, Michael Bynevelt, and Melinda Fitzgerald. Emerging Applications for Quantitative Susceptibility Mapping in the Detection of Traumatic Brain Injury Pathology. Neuroscience, 467:218–236, 2021.

[10] Christopher C. Giza and David A. Hovda. The new neurometabolic cascade of concussion. Neurosurgery, 75:S24–S33, 2014.

[11] Kendall R. Walker and Giuseppina Tesco. Molecular mechanisms of cognitive dysfunction following traumatic brain injury. Frontiers in Aging Neuroscience, 5, 2013.

[12] Daniel B. Hier, Tayo Obafemi-Ajayi, Matthew S. Thimgan, Gayla R. Olbricht, Sima Azizi, Blaine Allen, Bassam A. Hadi, and Donald C. Wunsch. Blood biomarkers for mild traumatic brain injury: a selective review of unresolved issues. Biomarker Research, 9(1), 2021.

[13] Ekaterina Lunkova, Guido I. Guberman, Alain Ptito, and Rajeet Singh Saluja. Noninvasive magnetic resonance imaging techniques in mild traumatic brain injury research and diagnosis. Human Brain Mapping, 42(16):5477–5494, 2021.

[14] Glen A. Cook and Jason S. Hawley. A review of mild traumatic brain injury diagnostics: Current perspectives, limitations, and emerging technology. Military Medicine, 179(10):1083–1089, 2014.

[15] Max Wintermark, Pina C. Sanelli, Yoshimi Anzai, A John Tsiouris, and Christopher T. Whitlow. Imaging evidence and recommendations for traumatic brain injury: Advanced neuro- and neurovascular imaging techniques. American Journal of Neuroradiology, 36(2):E1–E11, 2015.

[16] Liyan Lu, Heli Cao, Xiaoer Wei, Yuehua Li, and Wenbin Li. Iron Deposition Is Positively Related to Cognitive Impairment in Patients with Chronic Mild Traumatic Brain Injury: Assessment with Susceptibility Weighted Imaging. BioMed Research International, 2015, 2015.

[17] Eytan Raz, Jens H. Jensen, Yulin Ge, James S. Babb, L. Miles, Joesph Reaume, Robert I. Grossman, and Matilde Inglese. Brain iron quantification in mild traumatic brain injury: A magnetic field correlation study. American Journal of Neuroradiology, 32(10):1851–1856, 2011.

[18] Jeff H. Duyn and John Schenck. Contributions to magnetic susceptibility of brain tissue. NMR in Biomedicine, 30(4), 2017.

[19] Jinhee Jang, Yoonho Nam, Sung Won Jung, Tae Ryong Riew, Sang Hyun Kim, and In Beom Kim. Paradoxical paramagnetic calcifications in the globus pallidus: An ex vivo MR investigation and histological validation study. NMR in Biomedicine, 34(10), 2021.

[20] Sangwoo Kim, Youngjeon Lee, Chang Yeop Jeon, Keunil Kim, Youngjae Jeon, Yeung Bae Jin, Sukhoon Oh, and Chulhyun Lee. Quantitative magnetic susceptibility assessed by 7T magnetic resonance imaging in Alzheimer’s disease caused by streptozotocin administration. Quantitative Imaging in Medicine and Surgery, 10(3):789– 797, 2020.

[21] Yi Wang, Pascal Spincemaille, Zhe Liu, Alexey Dimov, Kofi Deh, Jianqi Li, Yan Zhang, Yihao Yao, Kelly M. Gillen, Alan H. Wilman, Ajay Gupta, Apostolos John Tsiouris, Ilhami Kovanlikaya, Gloria Chia Yi Chiang, Jonathan W. Weinsaft, Lawrence Tanenbaum, Weiwei Chen, Wenzhen Zhu, Shixin Chang, Min Lou, Brian H. Kopell, Michael G. Kaplitt, David Devos, Toshinori Hirai, Xuemei Huang, Yukunori Korogi, Alexander Shtilbans, Geon Ho Jahng, Daniel Pelletier, Susan A. Gauthier, David Pitt, Ashley I. Bush, Gary M. Brittenham, and Martin R. Prince. Clinical quantitative susceptibility mapping (QSM): Biometal imaging and its emerging roles in patient care. Journal of Magnetic Resonance Imaging, 46(4):951–971, 2017.

[22] Nan Jie Gong, Russell Dibb, Marjolein Bulk, Louise van der Weerd, and Chunlei Liu. Imaging beta amyloid aggregation and iron accumulation in Alzheimer’s disease using quantitative susceptibility mapping MRI. NeuroImage, 191:176–185, 2019.

[23] Zhiyong Zhao, Lei Zhang, Qingqing Wen, Wanrong Luo, Weihao Zheng, Tingting Liu, Yi Zhang, Keqing Zhu, and Dan Wu. The effect of beta-amyloid and tau protein aggregations on magnetic susceptibility of anterior hippocampal laminae in Alzheimer’s diseases. NeuroImage, 244, 2021.

[24] Christian Langkammer, Ferdinand Schweser, Nikolaus Krebs, Andreas Deistung, Walter Goessler, Eva Scheurer, Karsten Sommer, Gernot Reishofer, Kathrin Yen, Franz Fazekas, Stefan Ropele, and Jürgen R. Reichenbach. Quantitative susceptibility mapping (QSM) as a means to measure brain iron? A post mortem validation study. NeuroImage, 62(3):1593–1599, 2012.

[25] Chunlei Liu, Wei Li, Karen A. Tong, Kristen W. Yeom, and Samuel Kuzminski. Susceptibility-weighted imaging and quantitative susceptibility mapping in the brain. Journal of Magnetic Resonance Imaging, 42(1):23–41, 2015.

[26] Parsa Ravanfar, Samantha M. Loi, Warda T. Syeda, Tamsyn E. Van Rheenen, Ashley I. Bush, Patricia Desmond, Vanessa L. Cropley, Darius J.R. Lane, Carlos M. Opazo, Bradford A. Moffat, Dennis Velakoulis, and Christos Pantelis. Systematic Review: Quantitative Susceptibility Mapping (QSM) of Brain Iron Profile in Neurodegenerative Diseases. Frontiers in Neuroscience, 15, 2021.

[27] Suna Huang, Su Li, Hua Feng, and Yujie Chen. Iron Metabolism Disorders for Cognitive Dysfunction After Mild Traumatic Brain Injury. Frontiers in Neuroscience, 15, 2021.

[28] Eric J. Nisenbaum, Dmitry S. Novikov, and Yvonne W. Lui. The presence and role of iron in mild traumatic brain injury: An imaging perspective. Journal of Neurotrauma, 31:301–307, 2014.

[29] Sonia Levi, Maddalena Ripamonti, Andrea Stefano Moro, and Anna Cozzi. Iron imbalance in neurodegeneration. Molecular Psychiatry, 29(4):1139–1152, 2024.

[30] Maria Daglas and Paul A. Adlard. The Involvement of Iron in Traumatic Brain Injury and Neurodegenerative Disease. Frontiers in Neuroscience, 12, 2018.

[31] Sicheng Tang, Pan Gao, Hanmin Chen, Xiangyue Zhou, Yibo Ou, and Yue He. The Role of Iron, Its Metabolism and Ferroptosis in Traumatic Brain Injury. Frontiers in Cellular Neuroscience, 14, 2020.

[32] Edward Neuwelt, Joan Abbott, Lauren Abrey, William A Banks, Brian Blakley, Thomas Davis, Britta Engelhardt, Paula Grammas, Maiken Nedergaard, John Nutt, William Pardridge, Gary A Rosenberg, Quentin Smith, and Lester R Drewes. Strategies to advance translational research into brain barriers. The Lancet Neurology, 7(1):84–96, 2008.

[33] Maria Cristina Morganti-Kossmann, Laveniya Satgunaseelan, Nicole Bye, and Thomas Kossmann. Modulation of immune response by head injury. Injury, 38(12):1392–1400, 2007.

[34] Constantin Bouras, Panteleimon Giannakopoulos, Paul F. Good, Amy Hsu, Patrick R. Hof, and Daniel P. Perl. A Laser Microprobe Mass Analysis of Brain Aluminum and Iron in Dementia pugilistica: Comparison with Alzheimer’s Disease. Eur Neurol, 38(1):53–58, 1997.

[35] Luigi Zecca, Moussa B.H. Youdim, Peter Riederer, James R. Connor, and Robert R. Crichton. Iron, brain ageing and neurodegenerative disorders. Nature Reviews Neuroscience, 5(11):863–873, 2004.

[36] Tiffany K. Bell, Muhammad Ansari, Julie M. Joyce, Leah J. Mercier, David G. Gobbi, Richard Frayne, Chantel Debert, and Ashley D Harris. Quantitative susceptibility mapping in adults with persistent-post concussion symptoms after mild traumatic brain injury: An exploratory study. American Journal of Neuroradiology, 2024.

[37] Benjamin L. Brett, Kevin M. Koch, L. Tugan Muftuler, Matthew Budde, Michael A. Mccrea, and Timothy B. Meier. Association of Head Impact Exposure with White Matter Macrostructure and Microstructure Metrics. Journal of Neurotrauma, 38(4):474–484, 2021.

[38] Nan Jie Gong, Samuel Kuzminski, Michael Clark, Melissa Fraser, Mark Sundman, Kevin Guskiewicz, Jeffrey R. Petrella, and Chunlei Liu. Microstructural alterations of cortical and deep gray matter over a season of high school football revealed by diffusion kurtosis imaging. Neurobiology of Disease, 119:79–87, 2018.

[39] K. M. Koch, T. B. Meier, R. Karr, A. S. Nencka, L. T. Muftuler, and M. McCrea. Quantitative susceptibility mapping after sports-related concussion. American Journal of Neuroradiology, 39(7):1215–1221, 2018.

[40] Kevin M. Koch, Andrew S. Nencka, Brad Swearingen, Anne Bauer, Timothy B. Meier, and Michael McCrea. Acute Post-Concussive Assessments of Brain Tissue Magnetism Using Magnetic Resonance Imaging. Journal of Neurotrauma, 38(7):848–857, 2021.

[41] Najratun Nayem Pinky, Chantel T. Debert, Sean P. Dukelow, Brian W. Benson, Ashley D. Harris, Keith O. Yeates, Carolyn A. Emery, and Bradley G. Goodyear. Multimodal magnetic resonance imaging of youth sport-related concussion reveals acute changes in the cerebellum, basal ganglia, and corpus callosum that resolve with recovery. Frontiers in Human Neuroscience, 16, 2022.

[42] Alexander M. Weber, Anna Pukropski, Christian Kames, Michael Jarrett, Shiroy Dadachanji, Jack Taunton, David K.B. Li, and Alexander Rauscher. Pathological insights from quantitative susceptibility mapping and diffusion tensor imaging in ice hockey players pre and post-concussion. Frontiers in Neurology, 9, 2018.

[43] David K. Wright, Terence J. OBrien, and Sandy R. Shultz. Sub-acute Changes on MRI Measures of Cerebral Blood Flow and Venous Oxygen Saturation in Concussed Australian Rules Footballers. Sports Medicine - Open, 8(1), 2022.

[44] Robert Zivadinov, Paul Polak, Ferdinand Schweser, Niels Bergsland, Jesper Hagemeier, Michael G. Dwyer, Deepa P. Ramasamy, John G. Baker, John J. Leddy, and Barry S. Willer. Multimodal Imaging of Retired Professional Contact Sport Athletes Does Not Provide Evidence of Structural and Functional Brain Damage. Journal of Head Trauma Rehabilitation, 33(5):E24–E32, 2018.

[45] Christi A Essex, Jenna L Merenstein, Devon K Overson, Trong-Kha Truong, David J Madden, Mayan J Bedggood, Helen Murray, Samantha J Holdsworth, Ashley W Stewart, Catherine Morgan, Richard L M Faull, Patria Hume, Alice Theadom, and Mangor Pedersen. Distribution of paramagnetic and diamagnetic cortical substrates following mild Traumatic Brain Injury: A depth-and curvature-based quantitative susceptibility mapping study. medRxiv, 2024.

[46] Remika Mito, Mangor Pedersen, Heath Pardoe, Donna Parker, Robert E. Smith, Jillian Cameron, Ingrid E. Scheffer, Samuel F. Berkovic, David N. Vaughan, and Graeme D. Jackson. Exploring individual fixel-based white matter abnormalities in epilepsy. Brain Communications, 6(1), 2024.

[47] Juan F. Domínguez, Ashley Stewart, Alex Burmester, Hamed Akhlaghi, Kieran O’Brien, Steffen Bollmann, and Karen Caeyenberghs. Improving quantitative susceptibility mapping for the identification of traumatic brain injury neurodegeneration at the individual level. Zeitschrift fur Medizinische Physik, 2024.

[48] Mayan J. Bedggood, Christi A. Essex, Alice Theadom, Samantha J. Holdsworth, Richard L.M. Faull, and Mangor Pedersen. Individual-level analysis of MRI T2 relaxometry in mild traumatic brain injury: Possible indications of brain inflammation. NeuroImage: Clinical, 43, 2024.

[49] Arnaud Attyé, Félix Renard, Monica Baciu, Elise Roger, Laurent Lamalle, Patrick Dehail, Hélène Cassoudesalle, and Fernando Calamante. TractLearn: A geodesic learning framework for quantitative analysis of brain bundles. NeuroImage, 233, 2021.

[50] Adam Clemente, Arnaud Attyé, Félix Renard, Fernando Calamante, Alex Burmester, Phoebe Imms, Evelyn Deutscher, Hamed Akhlaghi, Paul Beech, Peter H Wilson, Govinda Poudel, Juan F Dominguez, and Karen Caeyenberghs. Individualised Profiling of White Matter Organisation in Moderate-to-Severe Traumatic Brain Injury Patients Using TractLearn: A Proof-of-Concept Study. Brain Research, 1806, 2023.

[51] Amy E. Jolly, Maria Balae, Adriana Azor, Daniel Friedland, Stefano Sandrone, Neil S.N. Graham, Karl Zimmerman, and David J. Sharp. Detecting axonal injury in individual patients after traumatic brain injury. Brain, 144(1):92–113, 2021.

[52] Phoebe Imms, Adam Clemente, Evelyn Deutscher, Ahmed M Radwan, Hamed Akhlaghi, Paul Beech, Peter H Wilson, Andrei Irimia, Govinda Poudel, Juan F Domínguez Duque, and Karen Caeyenberghs. Exploring personalized structural connectomics for moderate to severe traumatic brain injury. Network Neuroscience, 1(7):160–183, 2022.

[53] Alice Theadom, Natalie Hardaker, Charlotte Bray, Richard Siegert, Kevin Henshall, Katherine Forch, Kris Fernando, Doug King, Mark Fulcher, Sam Jewell, Nusratnaaz Shaikh, Renata Bastos Gottgtroy, and Patria Hume. The Brain Injury Screening Tool (BIST): Tool development, factor structure and validity. PLoS ONE, 16(2 February), 2021.

[54] Krzysztof J. Gorgolewski, Tibor Auer, Vince D. Calhoun, R. Cameron Craddock, Samir Das, Eugene P. Duff, Guillaume Flandin, Satrajit S. Ghosh, Tristan Glatard, Yaroslav O. Halchenko, Daniel A. Handwerker, Michael Hanke, David Keator, Xiangrui Li, Zachary Michael, Camille Maumet, B. Nolan Nichols, Thomas E. Nichols, John Pellman, Jean Baptiste Poline, Ariel Rokem, Gunnar Schaefer, Vanessa Sochat, William Triplett, Jessica A. Turner, Gaël Varoquaux, and Russell A. Poldrack. The brain imaging data structure, a format for organizing and describing outputs of neuroimaging experiments. Scientific Data, 3, 2016.

[55] Arnaud Boré, Samuel Guay, Christophe Bedetti, Steven Meisler, and Nick GuenTher. Dcm2Bids, August 2023.

[56] Xiangrui Li, Paul S. Morgan, John Ashburner, Jolinda Smith, and Christopher Rorden. The first step for neuroimaging data analysis: DICOM to NIfTI conversion. Journal of Neuroscience Methods, 264:47–56, 2016.

[57] Brian B. Avants, Nicholas J. Tustison, Gang Song, Philip A. Cook, Arno Klein, and James C. Gee. A reproducible evaluation of ANTs similarity metric performance in brain image registration. NeuroImage, 54(3):2033–2044, 2011.

[58] Nicholas J. Tustison, Brian B. Avants, Philip A. Cook, Yuanjie Zheng, Alexander Egan, Paul A. Yushkevich, and James C. Gee. N4ITK: Improved N3 bias correction. IEEE Transactions on Medical Imaging, 29(6):1310–1320, 2010.

[59] Bruce Fischl. FreeSurfer, 2012.

[60] Jenna L. Merenstein, Jiayi Zhao, Devon K. Overson, Trong Kha Truong, Kim G. Johnson, Allen W. Song, and David J. Madden. Depth- and curvature-based quantitative susceptibility mapping analyses of cortical iron in Alzheimers disease. Cerebral Cortex, 34(2), 2024.

[61] Barbara Dymerska, Korbinian Eckstein, Beata Bachrata, Bernard Siow, Siegfried Trattnig, Karin Shmueli, and Simon Daniel Robinson. Phase unwrapping with a rapid opensource minimum spanning tree algorithm (ROMEO). Magnetic Resonance in Medicine, 85(4):2294–2308, 2021.

[62] Christian Kames, Vanessa Wiggermann, and Alexander Rauscher. Rapid two-step dipole inversion for susceptibility mapping with sparsity priors. NeuroImage, 167:276– 283, 2018.

[63] Berkin Bilgic, Mauro Costagli, Kwok-Shing Chan, Jeff Duyn, Christian Langkammer, Jongho Lee, Xu Li, Chunlei Liu, José P Marques, Carlos Milovic, Simon Robinson, Ferdinand Schweser, Karin Shmueli, Pascal Spincemaille, Sina Straub, Peter Van Zijl, and Yi Wang. Recommended Implementation of Quantitative Susceptibility Mapping for Clinical Research in The Brain: A Consensus of the ISMRM Electro-Magnetic Tissue Properties Study Group. Technical report, 2023.

[64] Ashley W. Stewart and Stefan. Bollman. QSMxT/QSMxT, 2022.

[65] Ashley W. Stewart, Simon D. Robinson, Kieran OBrien, Jin Jin, Georg Widhalm, Gilbert Hangel, Angela Walls, Jonathan Goodwin, Korbinian Eckstein, Monique Tourell, Catherine Morgan, Aswin Narayanan, Markus Barth, and Steffen Bollmann. QSMxT: Robust masking and artifact reduction for quantitative susceptibility mapping. Magnetic Resonance in Medicine, 87(3):1289–1300, 2022.

[66] Mark Jenkinson, Christian F. Beckmann, Timothy E.J. Behrens, Mark W. Woolrich, and Stephen M. Smith. FSL. NeuroImage, 62(2):782–790, 2012.

[67] Stephen M. Smith, Mark Jenkinson, Mark W. Woolrich, Christian F. Beckmann, Timothy E.J. Behrens, Heidi Johansen-Berg, Peter R. Bannister, Marilena De Luca, Ivana Drobnjak, David E. Flitney, Rami K. Niazy, James Saunders, John Vickers, Yongyue Zhang, Nicola De Stefano, J. Michael Brady, and Paul M. Matthews. Advances in functional and structural MR image analysis and implementation as FSL. In NeuroImage, volume 23, 2004.

[68] Mark W. Woolrich, Saad Jbabdi, Brian Patenaude, Michael Chappell, Salima Makni, Timothy Behrens, Christian Beckmann, Mark Jenkinson, and Stephen M. Smith. Bayesian analysis of neuroimaging data in FSL. NeuroImage, 45(1), 2009.

[69] Stephen M. Smith. Fast robust automated brain extraction. Human Brain Mapping, 17(3):143–155, 2002.

[70] Douglas N. Greve and Bruce Fischl. Accurate and robust brain image alignment using boundary-based registration. NeuroImage, 48(1):63–72, 2009.

[71] Mark Jenkinson, Peter Bannister, Michael Brady, and Stephen Smith. Improved Optimization for the Robust and Accurate Linear Registration and Motion Correction of Brain Images. NeuroImage, 17(2):825–841, 2002.

[72] Mark Jenkinson and Stephen Smith. A global optimisation method for robust affine registration of brain images. Medical Image Analysis, 5:143–156, 2001.

[73] Jürgen R. Reichenbach. The future of susceptibility contrast for assessment of anatomy and function. NeuroImage, 62(2):1311–1315, 2012.

[74] Yixin Ma, Iain P. Bruce, Chun Hung Yeh, Jeffrey R. Petrella, Allen W. Song, and Trong Kha Truong. Column-based cortical depth analysis of the diffusion anisotropy and radiality in submillimeter whole-brain diffusion tensor imaging of the human cortical gray matter in vivo. NeuroImage, 270, 2023.

[75] Alessandro Daducci, Stephan Gerhard, Alessandra Griffa, Alia Lemkaddem, Leila Cammoun, Xavier Gigandet, Reto Meuli, Patric Hagmann, and Jean Philippe Thiran. The Connectome Mapper: An Open-Source Processing Pipeline to Map Connectomes with MRI. PLoS ONE, 7(12), 2012.

[76] Rahul S. Desikan, Florent Ségonne, Bruce Fischl, Brian T. Quinn, Bradford C. Dickerson, Deborah Blacker, Randy L. Buckner, Anders M. Dale, R. Paul Maguire, Bradley T. Hyman, Marilyn S. Albert, and Ronald J. Killiany. An automated labeling system for subdividing the human cerebral cortex on MRI scans into gyral based regions of interest. NeuroImage, 31(3):968–980, 2006.

[77] Donald J. Tournier, Robert Smith, David Raffelt, Rami Tabbara, Thijs Dhollander, Maximilian Pietsch, Daan Christiaens, Ben Jeurissen, Chun Hung Yeh, and Alan Connelly. MRtrix3: A fast, flexible and open software framework for medical image processing and visualisation. NeuroImage, 202, 2019.

[78] Miriam D. Waehnert, Juliane Dinse, Marcel Weiss, Markus N. Streicher, P. Waehnert, Stefan Geyer, Robert Turner, and Pierre-Louis Bazin. Anatomically motivated modeling of cortical laminae. NeuroImage, 93:210–220, 2014.

[79] Miriam D. Waehnert, Juliane Dinse, Andreas Schäfer, Stefan Geyer, Pierre Louis Bazin, Robert Turner, and Christine Lucas Tardif. A subject-specific framework for in vivo myeloarchitectonic analysis using high resolution quantitative MRI. NeuroImage, 125:94–107, 2016.

[80] Rudolph Pienaar, Bruce Fischl, Verne Caviness, N Makris, and Patricia E Grant. A Methodology for analyzing curvature in the developing brain from preterm to adult. International Journal of Imaging Systems and Technology, 18(1), 2008.

[81] Bruce Fischl, Anders M Dale, and Marcus E Raichle. Measuring the thickness of the human cerebral cortex from magnetic resonance images. PNAS, 97(20):11050– 11055, 2000.

[82] John W. Tukey. Exploratory Data Analysis. Addison-Wesley Publishing Company, 1977.

[83] Yoav Benjamini and Yosef Hochberg. Controlling the False Discovery Rate: A Practical and Powerful Approach to MultipleTesting. Journal of the Royal Statistical Society. Series B (Methodological), 57(1):289–300, 1995.

[84] Emily Kroshus, Bernice Garnett, Matt Hawrilenko, Christine M. Baugh, and Jerel P. Calzo. Concussion under-reporting and pressure from coaches, teammates, fans, and parents. Social Science and Medicine, 134:66–75, 2015.

[85] Michael McCrea, Timothy Meier, Daniel Huber, Alain Ptito, Erin Bigler, Chantel T. Debert, Geoff Manley, David Menon, Jen Kai Chen, Rachel Wall, Kathryn J. Schneider, and Thomas McAllister. Role of advanced neuroimaging, fluid biomarkers and genetic testing in the assessment of sport-related concussion: A systematic review. British Journal of Sports Medicine, 51(12):919–929, 2017.

[86] Martha E. Shenton, Haitham M. Hamoda, Jennifer S. Schneiderman, Sylvain Bouix, Olga Pasternak, Yogesh Rathi, M. A. Vu, Manoj P. Purohit, Karl Helmer, Inga Koerte, A Bence P. Lin, C. F. Westin, Ron Kikinis, Maciej Kubicki, Robert A. Stern, and Ross Zafonte. A review of magnetic resonance imaging and diffusion tensor imaging findings in mild traumatic brain injury. Brain Imaging and Behavior, 6(2):137–192, 2012.

[87] Hongyue Ma, Yan Dong, Yanhui Chu, Yanqin Guo, and Luxin Li. The mechanisms of ferroptosis and its role in alzheimers disease. Frontiers in Molecular Biosciences, 9, 2022.

[88] Christine Ghadery, Lukas Pirpamer, Edith Hofer, Christian Langkammer, Katja Petrovic, Marisa Loitfelder, Petra Schwingenschuh, Stephan Seiler, Marco Duering, Eric Jouvent, Helena Schmidt, Franz Fazekas, Jean Francois Mangin, Hugues Chabriat, Martin Dichgans, Stefan Ropele, and Reinhold Schmidt. R2* mapping for brain iron: Associations with cognition in normal aging. Neurobiology of Aging, 36(2):925–932, 2015.

[89] James Stankiewicz, S Scott Panter, Mohit Neema, Ashish Arora, Courtney E Batt, and Rohit Bakshi. Iron in Chronic Brain Disorders: Imaging and Neurotherapeutic Implications. Neurotherapeutics: The Journal of the American Society for Experimental NeuroTherapeutics, 4:371–386, 2007.

[90] A J Wills, G V Sawle, R R Guilbert, and A R J Curtis. Palatal tremor and cognitive decline in neuroferritinopathy. Journal of Neurology, Neurosurgery & Psychiatry, 73(1):91–92, 2002.

[91] Ann C. Mckee and Daniel H. Daneshvar. The neuropathology of traumatic brain injury. In Handbook of Clinical Neurology, volume 127, pages 45–66. Elsevier B.V., 2015.

[92] Helen C. Murray, Chelsie Osterman, Paige Bell, Luca Vinnell, and Maurice A. Curtis. Neuropathology in chronic traumatic encephalopathy: a systematic review of comparative post-mortem histology literature. Acta Neuropathologica Communications, 10(1), 2022.

[93] Ann C. McKee, Thor D. Stein, Bertrand R. Huber, John F. Crary, Kevin Bieniek, Dennis Dickson, Victor E. Alvarez, Jonathan D. Cherry, Kurt Farrell, Morgane Butler, Madeline Uretsky, Bobak Abdolmohammadi, Michael L. Alosco, Yorghos Tripodis, Jesse Mez, and Daniel H. Daneshvar. Chronic traumatic encephalopathy (CTE): criteria for neuropathological diagnosis and relationship to repetitive head impacts. Acta Neuropathologica, 145(4):371–394, 2023.

[94] Erin D. Bigler and William L. Maxwell. Neuropathology of mild traumatic brain injury: Relationship to neuroimaging findings. Brain Imaging and Behavior, 6(2):108–136, 2012.

[95] Rochelle A. Basil, Margaret L. Westwater, Martin Wiener, and James C. Thompson. A Causal Role of the Right Superior Temporal Sulcus in Emotion Recognition From Biological Motion. Open Mind, 2(1):26–36, 2017.

[96] Michael S. Beauchamp. The social mysteries of the superior temporal sulcus. Trends in Cognitive Sciences, 19(9):489–490, 2015.

[97] Ben Deen, Kami Koldewyn, Nancy Kanwisher, and Rebecca Saxe. Functional organization of social perception and cognition in the superior temporal sulcus. Cerebral Cortex, 25(11):4596–4609, 2015.

[98] Amber M. Leaver, Laurent Renier, Mark A. Chevillet, Susan Morgan, Hung J. Kim, and Josef P. Rauschecker. Dysregulation of Limbic and Auditory Networks in Tinnitus. Neuron, 69(1):33–43, 2011.

[99] Grit Hein and Robert T. Knight. Superior temporal sulcus - It’s my area: Or is it?, 12 2008.

[100] Marianne Dieterich and Thomas Brandt. Functional brain imaging of peripheral and central vestibular disorders. Brain, 131(10):2538–2552, 2008.

[101] Nicolas Boulloche, Marie Denuelle, Pierre Payoux, Nelly Fabre, Yves Trotter, and Gilles Géraud. Photophobia in migraine: An interictal PET study of cortical hyperexcitability and its modulation by pain. Journal of Neurology, Neurosurgery and Psychiatry, 81(9):978–984, 2010.

[102] Harvey S. Levin, Eugenio Amparo, Howard M. Eisenberg, David H. Williams, Walter M. High, Craig B. McArdle, and Richard L. Weiner. Magnetic resonance imaging and computerized tomography in relation to the neurobehavioral sequelae of mild and moderate head injuries. Journal of Neurosurgery, 66(5):706 – 713, 1987.

[103] Steven Kornguth, Neal Rutledge, Gabe Perlaza, James Bray, and Allen Hardin. A Proposed Mechanism for Development of CTE Following Concussive Events: Head Impact, Water Hammer Injury, Neurofilament Release, and Autoimmune Processes. Brain Sciences, 7(12):164, 2017.

[104] Jace B. King, Melissa P. Lopez-Larson, and Deborah A. Yurgelun-Todd. Mean cortical curvature reflects cytoarchitecture restructuring in mild traumatic brain injury. NeuroImage: Clinical, 11:81–89, 2016.

[105] Mazdak Ghajari, Peter J. Hellyer, and David J. Sharp. Computational modelling of traumatic brain injury predicts the location of chronic traumatic encephalopathy pathology. Brain, 140(2):333–343, 2017.

[106] Masaki Fukunaga, Tie Qiang Li, Peter Van Gelderen, Jacco A. De Zwart, Karin Shmueli, Bing Yao, Jongho Lee, Dragan Maric, Maria A. Aronova, Guofeng Zhang, Richard D. Leapman, John F. Schenck, Hellmut Merkle, and Jeff H. Duyn. Layer-specific variation of iron content in cerebral cortex as a source of MRI contrast. Proceedings of the National Academy of Sciences of the United States of America, 107(8):3834–3839, 2010.

[107] Hyeong Geol Shin, Jingu Lee, Young Hyun Yun, Seong Ho Yoo, Jinhee Jang, Se Hong Oh, Yoonho Nam, Sehoon Jung, Sunhye Kim, Masaki Fukunaga, Woojun Kim, Hyung Jin Choi, and Jongho Lee. χ-separation: Magnetic susceptibility source separation toward iron and myelin mapping in the brain. NeuroImage, 240, 2021.

[108] Yasushi Miyashita. Operating principles of the cerebral cortex as a six-layered network in primates: beyond the classic canonical circuit model. Proceedings of the Japan Academy Series B: Physical and Biological Sciences, 98(3):93–111, 2022.

[109] Lara Pankatz, Philine Rojczyk, Johanna Seitz-Holland, Sylvain Bouix, Leonard B. Jung, Tim L.T. Wiegand, Elena M. Bonke, Nico Sollmann, Elisabeth Kaufmann, Holly Carrington, Twishi Puri, Yogesh Rathi, Michael J. Coleman, Ofer Pasternak, Mark S. George, Thomas W. McAllister, Ross Zafonte, Murray B. Stein, Christine E. Marx, Martha E. Shenton, and Inga K. Koerte. Adverse Outcome Following Mild Traumatic Brain Injury Is Associated with Microstructure Alterations at the Gray and White Matter Boundary. Journal of Clinical Medicine, 12(16), 2023.

[110] Jun Liu, Zhi Feng Kou, and Yong Quan Tian. Diffuse axonal injury after traumatic cerebral microbleeds: An evaluation of imaging techniques, 2014.

[111] Ann C. McKee, Thor D. Stein, Christopher J. Nowinski, Robert A. Stern, Daniel H. Daneshvar, Victor E. Alvarez, Hyo Soon Lee, Garth Hall, Sydney M. Wojtowicz, Christine M. Baugh, David O. Riley, Caroline A. Kubilus, Kerry A. Cormier, Matthew A. Jacobs, Brett R. Martin, Carmela R. Abraham, Tsuneya Ikezu, Robert Ross Reichard, Benjamin L. Wolozin, Andrew E. Budson, Lee E. Goldstein, Neil W. Kowall, and Robert C. Cantu. The spectrum of disease in chronic traumatic encephalopathy. Brain, 136(1):43–64, 2013.

[112] Danielle K. Sandsmark, Asma Bashir, Cheryl L. Wellington, and R. Diaz-Arrastia. Cerebral Microvascular Injury: A Potentially Treatable Endophenotype of Traumatic Brain Injury-Induced Neurodegeneration. Neuron, 103(3):367–379, 2019.

[113] Yingxi Wu, Haijian Wu, Xinying Guo, Brock Pluimer, and Zhen Zhao. BloodBrain Barrier Dysfunction in Mild Traumatic Brain Injury: Evidence From Preclinical Murine Models. Frontiers in Physiology, 11, 2020.

[114] Roberta J. Ward, Fabio A. Zucca, Jeff H. Duyn, Robert R. Crichton, and Luigi Zecca. The role of iron in brain ageing and neurodegenerative disorders. The Lancet Neurology, 13(10):1045–1060, 2014.

[115] Rong Zhou, Bin Ji, Yanyan Kong, Limei Qin, Wuwei Ren, Yihui Guan, and Ruiqing Ni. PET Imaging of Neuroinflammation in Alzheimers Disease. Frontiers in Immunology, 12:739130, 2021.

[116] Jongho Lee, Sooyeon Ji, and Se Hong Oh. So You Want to Image Myelin Using MRI: Magnetic Susceptibility Source Separation for Myelin Imaging, 2024.

[117] Zhenghao Li, Ruimin Feng, Qiangqiang Liu, Jie Feng, Guoyan Lao, Ming Zhang, Jun Li, Yuyao Zhang, and Hongjiang Wei. APART-QSM: An improved sub-voxel quantitative susceptibility mapping for susceptibility source separation using an iterative data fitting method. NeuroImage, 274, 2023.

[118] Kathryn Wunderle, Kathleen M. Hoeger, Erin Wasserman, and Jeffrey J. Bazarian. Menstrual phase as predictor of outcome after mild traumatic brain injury in women. Journal of Head Trauma Rehabilitation, 29(5):E1–E8, 2014.

[119] Virginia Gallagher, Natalie Kramer, Kristin Abbott, John Alexander, Hans Breiter, Amy Herrold, Tory Lindley, Jeffrey Mjaanes, and James Reilly. The effects of sex differences and hormonal contraception on outcomes after collegiate sports-related concussion. Journal of Neurotrauma, 35(11):1242–1247, 2018.

[120] Ryan T. Tierney, Michael R. Sitler, C. Buz Swanik, Kathleen A. Swanik, Michael Higgins, and Joseph Torg. Gender differences in head-neck segmnet dynamic stabilization during head accelaration. Medicine and Science in Sports and Exercise, 37(2):272–279, 2005.

